# Identifying and ranking novel independent features for cardiovascular disease prediction in people with type 2 diabetes

**DOI:** 10.1101/2023.10.23.23297398

**Authors:** K Dziopa, N Chaturvedi, F W Asselbergs, A F Schmidt

## Abstract

**Background:** CVD prediction models do not perform well in people with diabetes. We therefore aimed to identify novel predictors for six facets of CVD, (including coronary heart disease (CHD), Ischemic stroke, heart failure (HF), and atrial fibrillation (AF)) in people with T2DM.

**Methods:** Analyses were conducted using the UK biobank and were stratified on history of CVD and of T2DM: 459,142 participants without diabetes or a history of CVD, 14,610 with diabetes but without CVD, and 4,432 with diabetes and a history of CVD. Replication was performed using a 20% hold-out set, ranking features on their permuted c-statistic.

**Results:** Out of the 600+ candidate features, we identified a subset of replicated features, ranging between 32 for CHD in people with diabetes to 184 for CVD+HF+AF in people without diabetes. Classical CVD risk factors (e.g. parental or maternal history of heart disease, or blood pressure) were relatively highly ranked for people without diabetes. The top predictors in the people with diabetes without a CVD history included: cystatin C, self-reported health satisfaction, biochemical measures of ill health (e.g. plasma albumin). For people with diabetes and a history of CVD top features were: self-reported ill health, and blood cell counts measurements (e.g. red cell distribution width). We additionally identified risk factors unique to people with diabetes, consisting of information on dietary patterns, mental health and biochemistry measures. Consideration of these novel features improved risk classification, for example per 1000 people with diabetes 133 CVD and 165 HF cases appropriately received a higher risk.

**Conclusion:** Through data-driven feature selection we identified a substantial number of features relevant for prediction of cardiovascular risk in people with diabetes, the majority of which related to non-classical risk factors such as mental health, general illness markers, and kidney disease.

## INTRODUCTION

We, and others ^1^ ^2^, have shown that cardiovascular (CVD) risk prediction models do not perform well in people with type 2 diabetes (T2DM). Importantly, performance did not differ meaningfully between 22 CVD risk prediction models, with c-statistics, estimating discrimination, close to 0.70 ^3^, while for general population this is 0.88 in women and 0.86 in men (based on the QRISK3^4^). This near-constant lower performance of most CVD prediction rules in people with T2DM likely reflect the considerable overlap in considered predictor variables, such as age, sex, blood pressure, and cholesterol reflecting a focus on features with a proven CVD association. The relative poor performance in people with T2DM identifies a need to consider less classical features for CVD prediction.

The need to include novel predictors for CVD has previously been shown by Wang *et. al.* ^5^, where up to 20% of patients with coronary disease did not possess conventional CVD risk factors, and 40% presented with only a single risk factor. Analysis strategies which *de-emphasize* model building, in favour of feature selection may therefore identify novel predictor variables, which is especially relevant for CVD prediction in people with T2DM where the currently available models do not perform well.

The UK Biobank (UKB) was initiated to further understanding of health in all its facets, and therefore collects measurements irrespective of clinical indication. For example, in clinical settings glucose and glycated haemoglobin (HbA_1c_) are typically only measured in people with, or at risk for, diabetes. In the UKB these features have been measured for nearly all enrolled participants, where during initial assessment information was collected on basic lifestyle and health information, anthropometric measurements, blood and urine samples, body composition, as well as a wealth of additional features. The large amounts of available measurements, taken independent of clinical indication, make the UKB particularly suited for a “hypothesis-free” data-driven approach to potentially uncover novel features.

The current study aimed to identify novel features for the 10-years risk of CVD in people with T2DM, which may be used to improve attempts at early identification of high-risk individuals and help with the management of CVD. We therefore crafted an integrated data engineering and feature selection pipeline to identify the subset of 600+ UKB measured feature which are predictive of the onset of CVD during a 10-year follow-up period. predictors for the 10 year risk of CVD, Analyses were conducted in three distinct groups of patients based on their clinical risk of CVD: without a history of diabetes or CVD at enrolment (T2DM/CVD), with T2DM at enrolment, and with T2DM&CVD at enrolment.

## METHODS

### Data source

Data was sourced from the UKB, a cohort of ∼500,000 men and women aged 40-69 years between 2006 and 2010 enrolled from primary care registers across the UK ^6^. These data were stratified into three groups: people without a diagnosis of CVD or T2DM at enrolment (wo T2DM/CVD), the second group included patients with T2DM diagnosis but no history of CVD at enrolment (w T2DM,), and the final group included individuals with diabetes and a history of CVD at enrolment (w T2DM&CVD). Follow-up considered the time from enrolment until the first CVD event, death or end of the study (10-years after enrolment), whichever came first. The candidate predictors were measured at the time of enrolment.

To identify novel features associating with the 10-years risk of CVD, we extracted variables (data fields) from 31 distinct UKB categories. The selected fields considered a range of information including anthropometry, blood chemistry, questionnaire data, and sociodemographic characteristics, jointly consisting of 603 unique variables; see Appendix Table 2. Please see Appendix Methods and Appendix Tables 3 – 7 for an overview of the sourced data and applied data engineering strategy.

### Statistical analysis

After randomly splitting the data into 80% for training, and 20% for testing, the training data were used to prune data on multicollinearity (Spearman’s correlation ≥ ±0.70) and absence of an outcome associations (univariable p-value ≥0.80).

To identify novel CVD-related features we leveraged a generalized linear model with a binomial distribution and an elastic net penalty^7^ (combining L1 and L2 regularisation), seamlessly removing less important features^7^. Ten-fold cross-validation, stratified by case (people who developed CVD) and control status (people who did not develop CVD), was used to optimize model hyper-parameters.

The feature importance of each selected variable was evaluated using a permutation feature importance algorithm (using 10 permutations) applied to the test data, quantifying the change in the c-statistic. Features were subsequently ranked by their c-statistic change, stratified by outcome type (CVD+AF+HF, CVD, CHD, HF, AF, Isch. Stroke) and a participant group (“wo T2DM/CVD”, “w T2DM”, “w T2DM&CVD”). By estimating the feature importance in the test data we were able to identify replicated findings (features with positive importance), which were unaffected by any potential overfitting. We dropped features with a zero or negative feature importance in the test data, indicating a failure to replicate. As age and sex are well-known and dominant CVD risk factors, the main text focussed on the remaining features, noting these remaining features are conditionally independent of age and sex; see full results in Appendix Tables 8 – 10 and Appendix Data 1.

Next, we sought to identify the importance of traditional CVD risk factors relative to our list of replicated novel risk factors. For this purpose we determined the rank of features used in any the following three clinically used prediction models: ASCVD^8^, QRISK3^4^ and the Framingham 1998^9^ score.

## RESULTS

### Patient characteristics

Data were available on 459,142 participants without T2DM and CVD at baseline “wo T2DM/CVD”, 14,610 individuals with T2DM but without a history of CVD at the time of diagnosis “w T2DM”, and 4,432 participants who had a history of CVD at the time of T2DM diagnosis “w T2DM&CVD”. Participants with CVD were on average older, male, had a higher BMI, and higher HbA_1c_ concentrations; see Table 1. During a median follow-up time of 10 years, 40,350 (8.8%) of the “wo T2DM/CVD” participants experienced a CVD+HF+AF event, with 2,671 (18.3%) CVD+HF+AF events in the “w T2DM” group, and 3,453 (77.9%) of the “w T2DM\&CVD” group; Appendix Table 11.

**Table 1.**
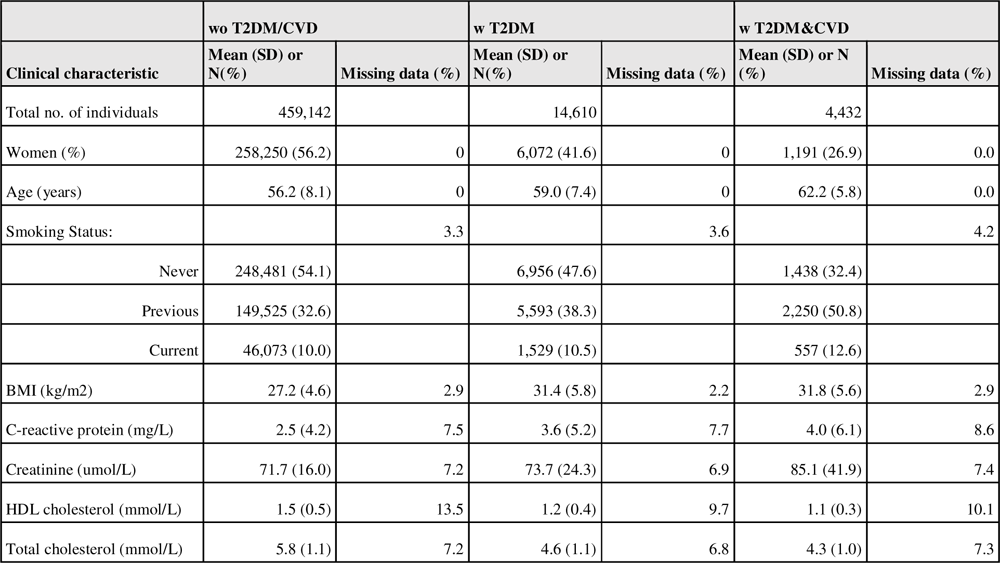
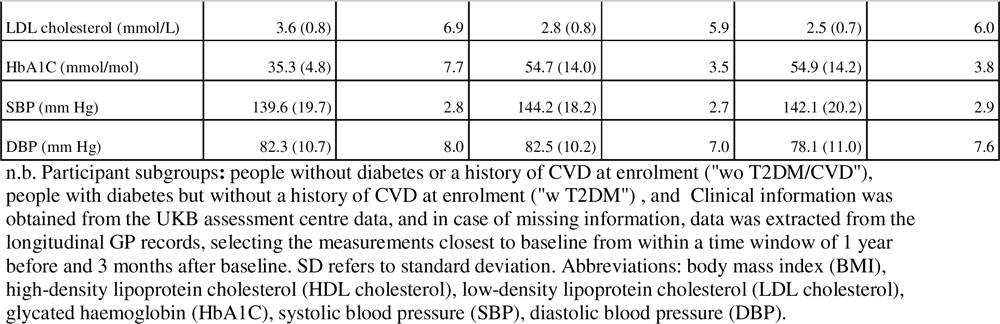
Clinical characteristics of UK biobank participants stratified by history of CVD and T2DM at enrolment.

|  | wo T2DM/CVD |  | w T2DM |  | w T2DM&CVD |  |
| --- | --- | --- | --- | --- | --- | --- |
| Clinical characteristic | Mean (SD) or N(%) | Missing data (%) | Mean (SD) or N(%) | Missing data (%) | Mean (SD) or N (%) | Missing data (%) |
| Total no. of individuals | 459,142 |  | 14,610 |  | 4,432 |  |
| Women (%) | 258,250 (56.2) | 0 | 6,072 (41.6) | 0 | 1,191 (26.9) | 0.0 |
| Age (years) | 56.2 (8.1) | 0 | 59.0 (7.4) | 0 | 62.2 (5.8) | 0.0 |
| Smoking Status: |  | 3.3 |  | 3.6 |  | 4.2 |
| Never | 248,481 (54.1) |  | 6,956 (47.6) |  | 1,438 (32.4) |  |
| Previous | 149,525 (32.6) |  | 5,593 (38.3) |  | 2,250 (50.8) |  |
| Current | 46,073 (10.0) |  | 1,529 (10.5) |  | 557 (12.6) |  |
| BMI (kg/m<sup>2</sup>) | 27.2 (4.6) | 2.9 | 31.4 (5.8) | 2.2 | 31.8 (5.6) | 2.9 |
| C-reactive protein (mg/L) | 2.5 (4.2) | 7.5 | 3.6 (5.2) | 7.7 | 4.0 (6.1) | 8.6 |
| Creatinine (umol/L) | 71.7 (16.0) | 7.2 | 73.7 (24.3) | 6.9 | 85.1 (41.9) | 7.4 |
| HDL cholesterol (mmol/L) | 1.5 (0.5) | 13.5 | 1.2 (0.4) | 9.7 | 1.1 (0.3) | 10.1 |
| Total cholesterol (mmol/L) | 5.8 (1.1) | 7.2 | 4.6 (1.1) | 6.8 | 4.3 (1.0) | 7.3 |
| LDL cholesterol (mmol/L) | 3.6 (0.8) | 6.9 | 2.8 (0.8) | 5.9 | 2.5 (0.7) | 6.0 |
| HbA1C (mmol/mol) | 35.3 (4.8) | 7.7 | 54.7 (14.0) | 3.5 | 54.9 (14.2) | 3.8 |
| SBP (mm Hg) | 139.6 (19.7) | 2.8 | 144.2 (18.2) | 2.7 | 142.1 (20.2) | 2.9 |
| DBP (mm Hg) | 82.3 (10.7) | 8.0 | 82.5 (10.2) | 7.0 | 78.1 (11.0) | 7.6 |

n.b. Participant subgroups: people without diabetes or a history of CVD at enrolment ("wo T2DM/CVD"), people with diabetes but without a history of CVD at enrolment ("w T2DM") , and Clinical information was obtained from the UKB assessment centre data, and in case of missing information, data was extracted from the longitudinal GP records, selecting the measurements closest to baseline from within a time window of 1 year before and 3 months after baseline. SD refers to standard deviation. Abbreviations: body mass index (BMI), high-density lipoprotein cholesterol (HDL cholesterol), low-density lipoprotein cholesterol (LDL cholesterol), glycated haemoglobin (HbA1C), systolic blood pressure (SBP), diastolic blood pressure (DBP).

### Prioritized features for CVD prediction

Out of the 603 initially available UKB data fields, 382 were retained after the data engineering steps, depending on the type of CVD and patient subgroup between 229 and 258 features remainder after filtering on univariable association and multicollinearity; see Appendix Table 12, Figure 1. An elastic net algorithm was applied to identify a subset of variables associated with CVD outcomes, which were subsequently replicated in the independent test set, resulting in a range of replicated features between 32 (for CHD in “w T2DM”) and 200 (for Isch. Stroke in “w T2DM&CVD”); Figure 1. Generally, our pipeline identified the most features for the wo T2DM/CVD subgroup (on average 156 features were replicated across the six considered CVD outcomes), followed by w T2DM&CVD (an average of 126 features),, and w T2DM (an average of 63 features); Figure 1.

**Figure 1.**
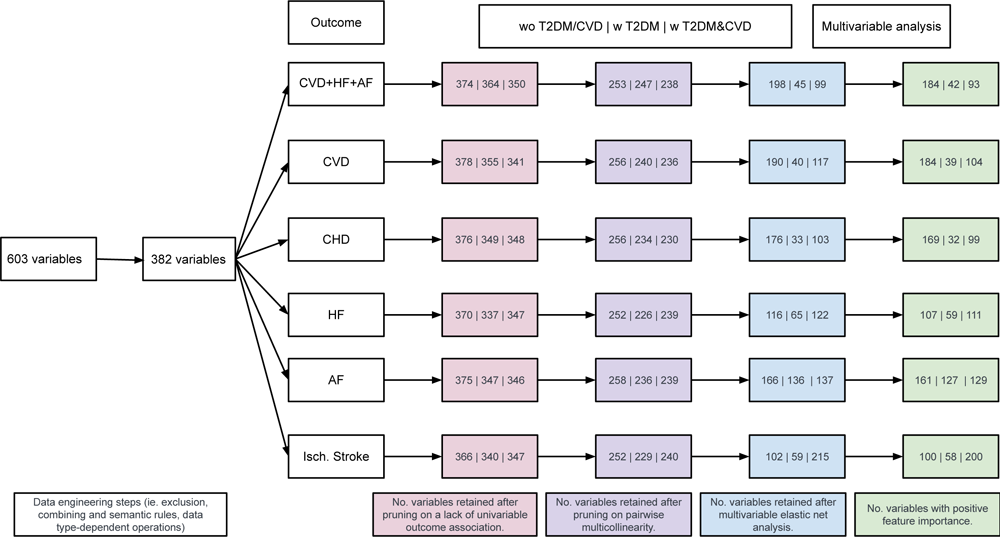
Overview of features available at the different stages of our feature selection algorithm. n. b. Variables were removed based on an insufficient univariable association with the outcome (Spearman’s p-value greater than 0.8), because of a high pairwise correlation (Spearman’s correlation above 0.70), through elastic net based penalization, or because of a negative feature importance in the testing data (representing independent replication). Abbreviations: type 2 diabetes (T2DM); cardiovascular disease (CVD); coronary heart disease (CHD); atrial fibrillation (AF); heart failure (HF); ischemic stroke (Isch. Stroke).

Ranking features on their summed c-statistic, aggregated across CVD outcomes, (Figure 2, Appendix Data 1, Appendix Tables 8 - 10) highlighted the importance of plasma biomarkers such as cystatin C, red blood cell distribution width (RDW), HbA_1c_, plasma albumin, plasma urate, testosterone, and urine microalbumin, as well as clinical characteristics such as diastolic and systolic blood pressure (DBP/SBP), and estimated trunk mass. Additionally, many of the top ranking features included indicators of a poor health status (e.g., “self-reported: health satisfaction”, “quit smoking due to illness”, “self-reported: recent tiredness”, “disability parking permit (blue badge)”), family (father, mother, sibling) history of heart disease. While systolic/diastolic blood pressure (SBP/DBP) were ranked 3^rd^ and 9^th^ respectively, lipids biomarkers conveyed relatively limited discriminative ability: low-density lipoprotein (LDL-C) was ranked 54^th^ and high-density lipoprotein (HDL-C) 37^th^.

**Figure 2.**
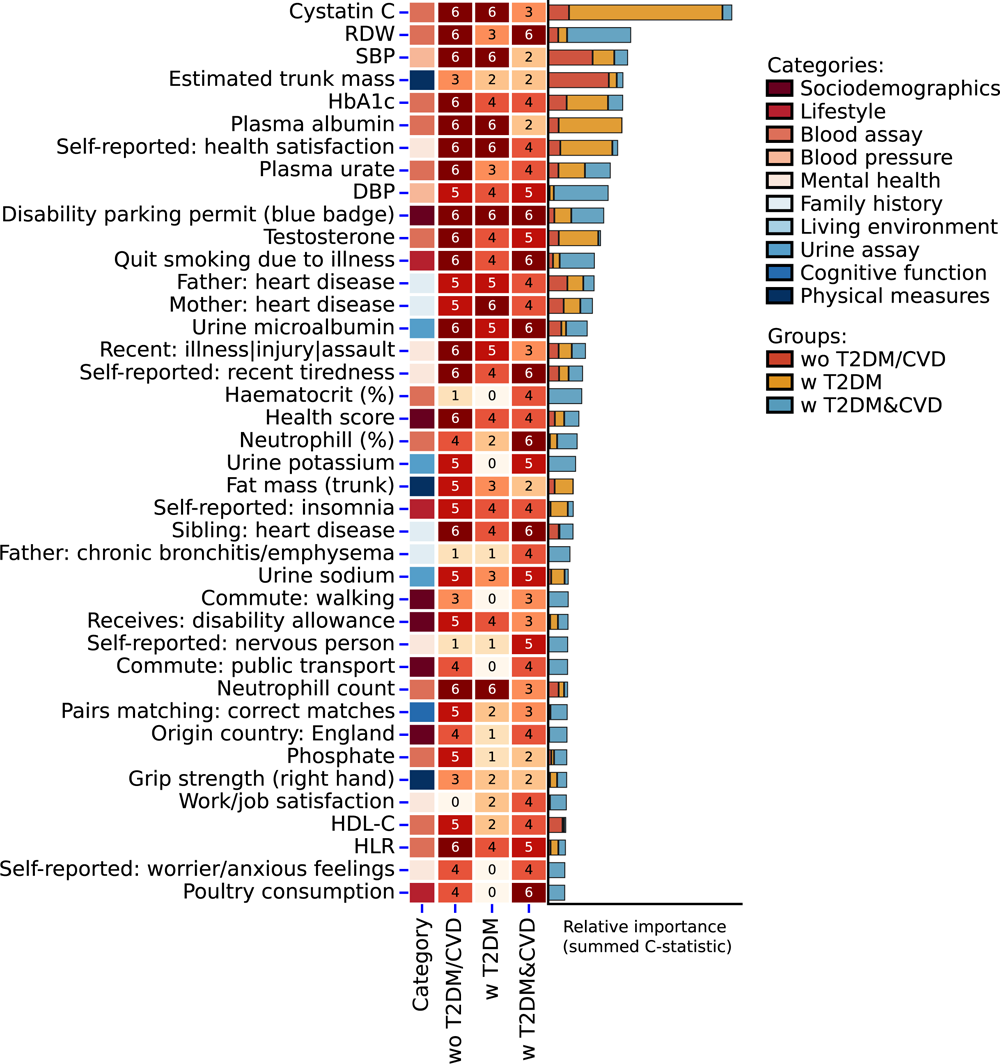
Top 40 most predictive features for the 10-years risk of cardiovascular disease n. b. The y-axis presents the top 40 features (excluding age and sex) based on the summed feature importance aggregated across the six types of CVD considered (CVD+AF+HF, CVD, CHD, Isch. Stroke, HF, AF) stratified by participants subgroup: people without diabetes or a history of CVD at enrolment (“wo T2DM/CVD”), people with diabetes but without a history of CVD at enrolment (“w T2DM”), and people with a history of diabetes and CVD at enrolment (“w T2DM\&CVD”). The heatmap (mid panel) represents the number of CVD outcomes the features was identified for (at most 6), while the stacked bar chart (right panel) encodes the summed feature importance, stratified by patient subgroup. Feature importance was calculated using a permuted feature importance algorithm recording the change in c-statistic. The algorithm was applied to the hold-out test data and hence represent an unbiased estimates of the feature importance as well as reflecting features which were independently replicated. The complete list of identified features is provided in Appendix Data 1. Abbreviations: type 2 diabetes (T2DM), cardiovascular disease (CVD), red blood cell distribution width (RDW), glycated haemoglobin (HbA1c), low-density lipoprotein cholesterol (LDL-C), gamma glutamyltransferase (GGT), sex hormone-binding globulin (SHBG), high light scatter reticulocyte count (HLR).

### The top 5 most relevant features per CVD type and participant subgroup

The top 5 most important features per CVD type and participant subgroup is presented in Figure 3, with the full list of features presented in Appendix Data 1, and Appendix Figures 4 – 6, and Appendix Tables 8 – 10. For people without T2DM or CVD at enrolment (wo T2DM/CVD), SBP and family history of heart disease were important features for CVD+AF+HF, CVD, and CHD, while HDL-C and HbA_1c_were particularly important for CVD and CHD. SBP, Cystatin C, and urine microalbumin populated the top 5 most important predictors for ischaemic stroke as well as HF, while estimated trunk mass was ranked highest for AF and HF; Figure 3.

**Figure 3.**
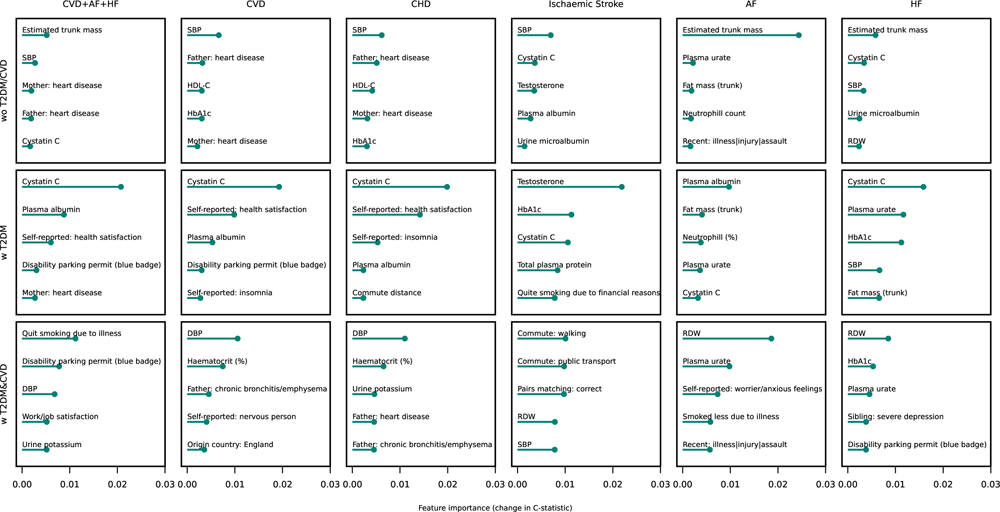
Top five most predictive features for the 10-years risk of cardiovascular disease, stratified by disease type. n.b. Results are stratified by type of CVD and participant subgroup: people without diabetes or a history of CVD at enrolment (“wo T2DM/CVD”), people with diabetes but without a history of CVD at enrolment (“w T2DM”), and people with a history of diabetes and CVD at enrolment (“w T2DM&CVD”). Feature importance was calculated using a permuted feature importance algorithm recording the change in c-statistic. The algorithm was applied to the hold-out test data and hence represent an unbiased estimates of the feature importance as well as reflecting features which were independently replicated. Abbreviations: type 2 diabetes (T2DM); cardiovascular disease (CVD); coronary heart disease (CHD); atrial fibrillation (AF); heart failure (HF); ischemic stroke (Isch. Stroke). systolic blood pressure (SBP), high-density lipoprotein cholesterol (HDL-C), glycated haemoglobin (HbA1c), red blood cell distribution width (RDW), diastolic blood pressure (DBP), disability parking permit (Receives: blue badge).

**Figure 4.**
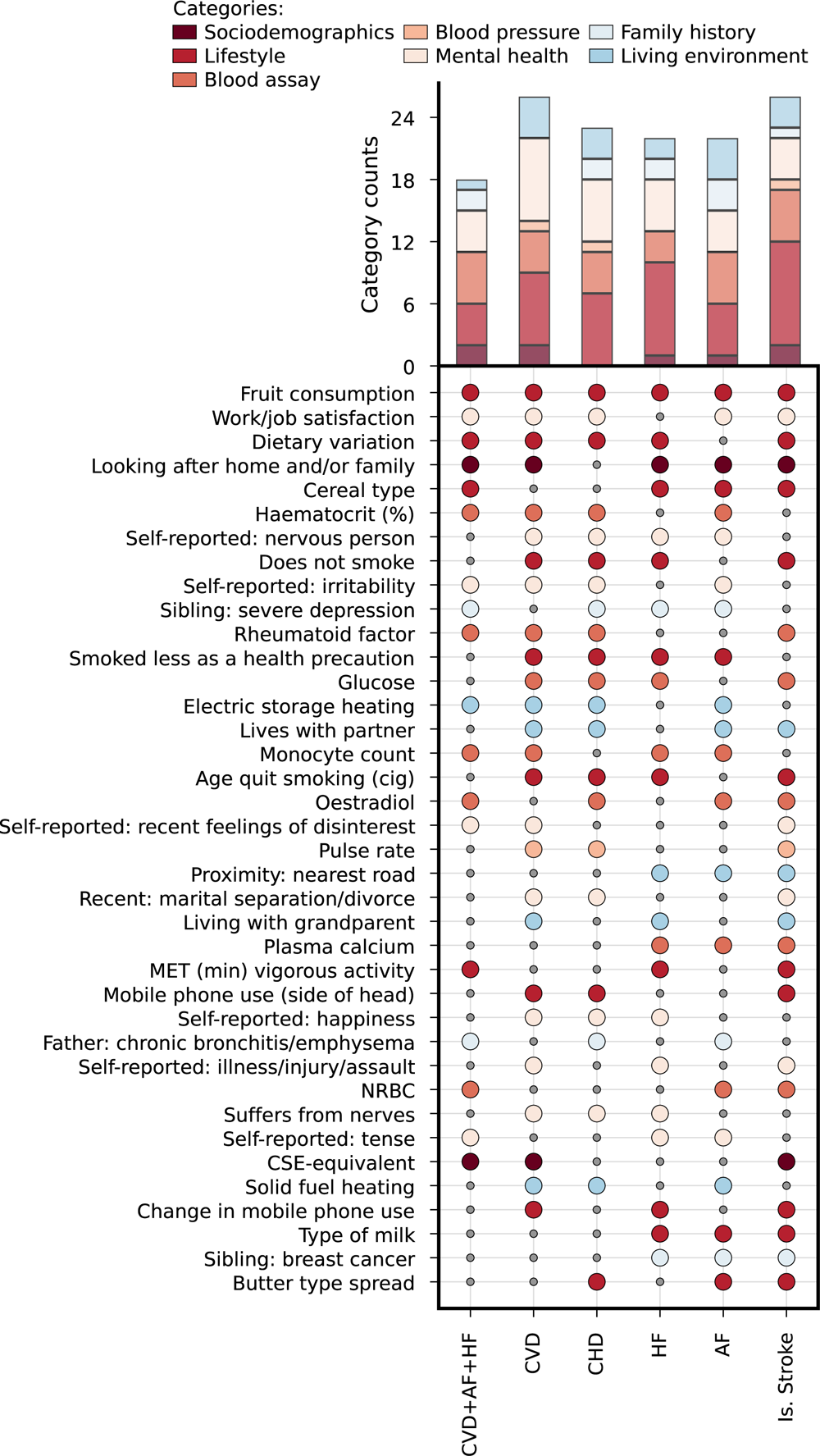
Diabetes specific features for the 10-years risk of cardiovascular disease N.B. The figure focuses on features which were predictive for at least three of the six considered CVD outcomes, the complete list of features is presented in Appendix Data 3 - 5. Abbreviations: type 2 diabetes (T2DM); cardiovascular disease (CVD); coronary heart disease (CHD); atrial fibrillation (AF); heart failure (HF); ischemic stroke (Isch. Stroke); metabolic equivalent task (MET), nucleated red blood cells (NRBC), qualifications (CSE-equivalent). The top panel reflect the number of times feature belonging to a specific UK biobank category was selected.

For people with T2DM at the time of enrolment (w T2DM), Cystatin C was particularly important to predict all 6 CVD outcomes, with a feature importance of 0.020 for CHD in the “w T2DM” group, compared to only 0.001 in people without diabetes. Self-reported health satisfaction and self-reported insomnia were important for CVD and CHD, where self-reported insomnia was also included as a top 5 predictor for CHD. HbA_1c_ was a strong predictor for Ischaemic stroke and HF, while fat mass and plasma urate were important for AF and HF; Figure 3 and Appendix Table 9.

For people with T2DM and CVD at the time of enrolment (w T2DM&CVD), indicators of, sometimes recent, adversity in (perceived) health or stress were important risk factors for CVD. For example, recent illness, injury or assault (AF), owning a disability parking permit (CVD+HF+AF, HF), self-reported nervousness (CVD), or self-reported worrier/anxious feelings (AF)Furthermore, familial history of disease was often included in the top 5: father with a history of chronic bronchitis/emphysema (CVD, CHD), father with a history of heart disease (CHD), sibling with a history of severe depression (HF). More traditional biomarkers/clinical measurements were also retained in the top 5: DBP (CHD+HF+AF, CVD, CHD), and SBP (ischaemic stroke), haematocrit (CVD, CHD), RDW (ischaemic stroke, AF, HF), plasma urate (AF, HF), and HbA_1c_ (HF); see Figure 3 and Appendix Table 10.

Finally, as detailed in Appendix Tables 8 - 10, we note that while age and sex were often the most important predictors irrespective of a participants diabetes status, in people with diabetes the c-statistic was severely attenuated compared to people without diabetes. For example, for CVD the c-statistic for age was 0.086 in the wo T2DM/CVD group, compared to 0.041 in the w T2DM group.

### CVD features selected in all three participants groups

We next identified features that were selected for all three participants subgroups, stratifying by CVD outcome type; Appendix Data 2 and Appendix Results. The number of common features ranged from 14 for CHD to 44 for AF. Briefly, we observed that HbA1c was an important predictor irrespective of the diabetes status, ranking highly for predictions of HF, AF, and Is. stroke. Interestingly, HbA1c was the 5^th^ most important predictor for CHD in people without diabetes, for people with diabetes glucose was selected instead of HbA1c for CHD prediction. Cystatin C and RDW were predictive of HF, AF and Is. stroke (cystatin C only) irrespective of the participant subgroup. Aside from these biochemistry measures, we observed that information on familial disease history, self-reported health (satisfaction), mental-health and socio-economic factors were often predictive of CVD irrespective of the diabetes status.

### CVD features unique for people with T2DM

Given the poor performance of CVD prediction models in people with T2DM, we next identified the union of features which were uniquely selected for “w T2DM” or “w T2DM&CVD” participant subgroups; Figure 4, Table 2, Appendix Data 3-4. On average 7 life style factor were unique to CVD prediction in people with diabetes, were particularly diet related information such as fruit consumption, dietary variability, or type of cereal was paramount. Information on mental health (on average 5 features) and blood assay (on average 4 features) were also important for CVD prediction in people with diabetes. For example, self-reported nervousness, irritability, recent feelings of disinterest, and recent divorce or separation were all relevant and unique to CVD prediction in people with diabetes. Similarly, plasma glucose, rheumatoid factor, haematocrit percentage, monocyte counts, and oestradiol were important and unique predictors for CVD in people with diabetes (Figure 4).

**Table 2.** The number of features for the 10-years risk of cardiovascular disease unique to people with diabetes, stratified by disease type.

|  | CVD+AF+HF | CVD | CHD | HF | AF | Is. Stroke |
| --- | --- | --- | --- | --- | --- | --- |
| **Features specific to w T2DM** | 0 | 2 | 3 | 13 | 21 | 6 |
| **Features specific to w T2DM&CVD** | 26 | 35 | 30 | 44 | 29 | 97 |
| **Common features for T2DM w/wo CVD** | 2 | 0 | 2 | 10 | 19 | 20 |
| **Union of features for T2DM w/wo CVD** | 113 | 121 | 115 | 140 | 193 | 207 |

n.b. Participant subgroups: people without diabetes or a history of CVD at enrolment ("wo T2DM/CVD"), people with diabetes but without a history of CVD at enrolment ("w T2DM") , and people with a history of diabetes and CVD at enrolment ("w T2DM&CVD"). Rows: "Features specific to w T2DM" – the number of unique features selected for "w T2DM" group excluding features selected for other groups; "Features specific to "w T2DM&CVD" – the number of unique features selected for "w T2DM&CVD" group excluding features selected for other two groups; "Common features for T2DM w/wo CVD" – the number of common features selected for "w T2DM" and "w T2DM&CVD" excluding features identified for "wo T2DM/CVD"; "Union of features for T2DM w/wo CVD" – the number of all unique features identified for "w T2DM" and "w T2DM&CVD" excluding features selected for "wo T2DM/CVD". Outcomes: cardiovascular disease including heart failure and/or atrial fibrillation (CVD+AF+HF), cardiovascular disease (CVD), coronary heart disease (CHD), heart failure (HF), atrial fibrillation (AF), ischaemic stroke (Is. Stroke).

### Ranking features used by three clinical CVD prediction models

We determined the importance and rank of 18 features included in at least one of the clinically used prediction models: ASCVD^8^, QRISK3^4^, and the Framingham^9^, Appendix Tables 13-15. In people without diabetes (“wo T2DM/CVD”) 9 features were selected to predict CVD, with the 7 in the top 10% (in order of importance: age, sex, SBP, paternal history of CVD, HDL-C, maternal history of CVD, and sibling history of CVD); Appendix Table 13. For the “w T2DM” participants, 8 features were selected for CVD, and only age and sex were retained in the top 10 most important features; Appendix Table 14. Similarly, for “w T2DM&CVD” participants 8 features were selected for CVD, where age, DBP and sex were the top 10 most important features; Appendix Tabel 15. The following features included in these 3 clinically used models were not selected in any of the three participant groups when predicting CVD: BMI, smoking status, total cholesterol, deprivation, moderate or severe depression, bipolar disorder. Finally, LDL-C was ranked 30.16% for people without diabetes, and not selected for people with diabetes irrespective of CVD history at enrolment.

### Clinical benefit of considering novel risk factors

We additionally estimated how many cases were appropriately assigned a higher risk by additionally considering information from the here identified non-classical risk factors. For this we compared risk group assignment (using the canonical risk groups <10%, between 10% and 20%, and ≥20%) based on the 18 classical risk factors used in the ASCVD^8^, QRISK3^4^, or the Framingham^9^ against risk group assignment combining classical and non-classical risk factors. Per 1,000 people without diabetes (the “wo T2DM/CVD” group)253 participants who went on to develop CVD appropriately received a higher risk, for HF this was 36. Per 1,000 people with diabetes (the “w T2DM” group) these numbers were 133 for CVD and 165 for HF.

## DISCUSSION

In this study, we leveraged data from the richly phenotyped UKB to identify potential novel predictors for CVD, specifically focusing on people with diabetes. Combining a bespoke data-engineering strategy with supervised feature selection using an elastic net algorithm, we found a prioritized list of 32 to 200 features, depending on the type of CVD considered. The classical risk factors (e.g. parental or maternal history of heart disease, and blood pressure) were relatively highly ranked for people without diabetes (“wo T2DM/CVD”), however, truncal mass was selected instead of BMI, and HDL-C rather than total cholesterol or LDL-C.

These traditional predictors were much less important in people with diabetes. Instead the following features were import to predict CVD in people with diabetes but without a history of CVD (“w T2DM”): HbA1c, cystatin C, self-reported health satisfaction, biochemical measures of ill health e.g. plasma albumin (representing liver and kidney damage). In people with diabetes and a history with CVD important features to predict CVD consisted of: self-reported ill health, and biochemical measures of ill health (RDW, haematocrit – representing anaemia) were selected. We additionally identified features common between people with and without diabetes. For example, HbA1c was important for people with and without diabetes to predict HF, AF, and Is. stroke, interestingly HbA1c was important for CHD prediction (ranked 5^th^) for people without diabetes. Similarly, cystatin C and RDW were a common predictor for HF, AF and Is. stroke. Focusing on features unique to people with diabetes highlighted the importance of information on glycaemic traits (e.g., plasma glucose concentration), diet (e.g. fruit consumption), physical activity, mental health (e.g., irritability, happiness, feelings of disinterest), socioeconomic status (e.g., work/job satisfaction), environment (e.g., proximity to nearest road), as well as plasma measurements of blood cells, calcium concentration, total protein, oestradiol, and rheumatoid factor.

Due to the unique design of the UK Biobank, where measurements are obtained from all participants, irrespective of potential clinical diagnosis, we were able to highlight the relevance of kidney and diabetes markers for CVD prediction in people without such an indication. Importantly, these features were typically more discriminative than lipid measurements, suggesting that currently available risk prediction tools for CVD might be further optimized by adding early markers for kidney disease and diabetes. This held true for people with and people without diabetes, for example HbA1c was in the top 5% of most important CHD predictors for people without diabetes. The observation that LDL-C is not strongly predictive of CHD has been reported before^10^ (popular risk scores such as QRISK3 ^4^ and Framingham^9^ do not consider LDL-C as a required predictor), and also illustrated by most CVD models instead including HDL-C or TG (out of 22 CVD risk prediction models considered by Dziopa *et al.* 2021, only a single model did not include HDL-C^3^). This of course does not imply that LDL-C does not have a causal effect on CHD or CVD, instead this highlights that good predictors of disease risk need not have a causal effect on disease manifestation. This adage is also exemplified by the selection algorithm including features which are (relatively) immutable, such as sex, maternal and paternal disease history, educational attainment or socioeconomic status.

While some of the here reported features have previously been associated with CVD and its individual components such as CHD, ischemic stroke, AF and HF, the current study is able to uniquely account for their pairwise correlation, thereby ensuring the identified features provide *independent* information. Additionally, by training a multivariable model we were able to estimate relative feature importance, and thereby rank features on their relevance for disease classification. Such a rank provides directly actionable information for clinicians and researchers wishing to enrich the currently available prediction algorithms. Furthermore, by identifying features for six types of CVD, we observe that many of the identified features are shared by distinct diseases such as CHD, HF, and AF. The commonality between features suggests that the same or similar information can be used to predict multiple types of CVD and hence better inform patient risk and optimize care, further supporting our previous study which generalized CVD risk prediction tools to predict the 10-years risk of HF and AF^3^.

This study has some limitations which warrant discussion. Firstly, the UKB predominantly consists of white European participants, with an on average higher socioeconomic status, and hence generalizability to other ethnicities should be explored. Secondly, some of the identified features might be specific to the UK, for example, a Blue Badge is the UK’s version of a disabled parking permit. We do expect that many of these UK-specific features can be implemented in different countries given sufficiently careful mapping, similar to how laboratory measurement might need to be recalibrated between distinct labs. Thirdly, we emphasize that the identified features, feature importance, and feature rank were based on the testing data and therefore are unaffected by any potential model overfit and at the same time represents independently replicated findings.

Nevertheless, with increasing sample size additional features will likely be identified, particularly for relatively infrequent outcomes such as Is. Stroke. Aside from the influence of sample size, due to the multivariate nature (where features are correlated among themselves), similar but slightly different, features might be identified in subsequent studies (e.g., SBP may stand in for DBP due to the correlation between both measurements). Due to the scale of our analyses, we have not considered exploring potential non-linear associations. While non-linear associations are to be expected in health and healthcare, often simplified models such as applied here are sufficient to detect the presence of an association, with subsequent research considering the possibility of non-linearity. For example, alcohol intake and LDL-C are known risk factors for CVD which, after initial discovery, are now thought to elicit a non-linear U-shaped association with disease onset.

## CONCLUSIONS

In this scaled analysis of the UK Biobank, we showed that the classical risk factors were less importance in people with diabetes. We have identified numerous independent variables which predict CVD in people with diabetes, covering information on mental health, familial CVD and non-CVD disease histories, as well as markers for general ill-health, early kidney disease and diabetes (control). We note that some of the identified features, such as HbA1c and cystatin C, predict CVD irrespective of the diabetes status. Furthermore, we identified diabetes specific features predicting CVD typically covering dietary patterns, mental health and biochemistry measures. The identified features are typically overlooked by currently available CVD prediction models and provide actionable leads to improve CVD prediction models.

## Data Availability

All data produced in the present study are available upon request to the UK Biobank.

## Conflict of interest statement

NC serves on data safety and monitoring committees of clinical trials sponsored by AstraZeneca. AFS has received funding from New Amsterdam Pharma for unrelated work. None of the other authors of this paper has a financial or personal relationship with other people or organizations that could inappropriately influence or bias the content of the paper.

## Author contributions

AFS, FWA, NC contributed to the idea and design of the study. KD prepared the dataset for analysis and implemented feature selection methods. KD and AFS conducted the data analysis and created the figures. KD wrote the manuscript with support from FWA, AFS, and NC. FWA, NC and AFS provided critical feedback on the analysis and its interpretation, and commented on the drafted manuscript. KD is responsible for the integrity of the work as a whole.

## Code availability

Analyses were carried out in Python v3.6 using *scikit-learn*^11^, *statsmodels*^12^, *pandas*^13^, and *numpy*^14^, plots were generated using *matplotlib*^15^, and *seaborn*^16^, imputation was performed in R v4.1 using *mice*^17^. See https://gitlab.com/cvd_in_t2dm/novel_features_cvd_prediction for the codebase underpinning this work.

## Acknowledgments

This research has been conducted using the UK Biobank Resource under application numbers 12113, 24711 and 44972. We are grateful to the UK Biobank participants. UK Biobank was established by the Wellcome Trust medical charity, Medical Research Council, Department of Health, Scottish Government, and the Northwest Regional Development Agency. It has also had funding from the Welsh Assembly Government and the British Heart Foundation.

## Funding and role of funding sources

KD is supported by a PhD studentship from the National Productivity Investment Fund – MRC Doctoral Training Programme (grant no. MR/S502522/1). AFS is supported by BHF grants PG/18/5033837, PG/22/10989, and the UCL BHF Research Accelerator AA/18/6/34223. AFS received additional support from the National Institute for Health Research University College London Hospitals Biomedical Research Centre. This work was supported by grant [R01 LM010098] from the National Institutes of Health (USA) and by EU/EFPIA Innovative Medicines Initiative 2 Joint Undertaking BigData@Heart grant n° 116074, as well as by the UKRI/NIHR Multimorbidity fund Mechanism and Therapeutics Research Collaborative MR/V033867/1 and the Rosetrees Trust. This work is partially supported by Dutch Research Council (628.011.213). This publication is part of the project MyDigiTwin with project number 628.011.213 of the research programme “COMMIT2DATA – Big Data & Health” which is partly financed by the Dutch Research Council (NWO).

## Prior postings and presentations

This study and its results have not been published previously. A preprint version has been deposited on medrxiv.

## ACRONYMS

Acronym: Meaning

AF: Atrial fibrillation

BMI: Body Mass Index

CHD: Coronary heart disease

CVD: Cardiovascular disease

CVD+HF+AF: Cardiovascular disease including heart failure and/or atrial fibrillation

DBP: Diastolic blood pressure

HDL-C: High-density lipoprotein cholesterol

HF: Heart failure

LDL-C: Low-density lipoprotein cholesterol

MI: Myocardial infarction

SBP: Systolic blood pressure

T2DM: Type 2 diabetes

UKB: UK Biobank

w T2DM: Individuals with T2DM diagnosis but not history of CVD

w T2DM&CVD: Individuals with T2DM and a history of CVD

wo T2DM/CVD: Individuals with T2DM and a history of CVD

## Notes

### Summary of Updates

Formatting abstract

