## Appendix for "Identifying and ranking novel independent features for cardiovascular disease prediction in people with type 2 diabetes"

October 20, 2023

### Appendix

#### 1.1 Methods

##### 1.1.1 Outcome definitions

CVD was defined as the occurrence of fatal or non-fatal myocardial infarction (MI), sudden cardiac death, ischemic heart disease, fatal or non-fatal stroke or PAD after the start of follow-up. We additionally considered a broader definition of CVD, also including heart failure (HF) and/or atrial fibrillation (AF): ”CVD+HF+AF”, as well as the individual CVD components: CHD, stroke, AF, and HF; see Appendix Table 1.

##### 1.1.2 UK Biobank structure

The data fields in the UK Biobank cover multiple time points, and within each time point, a variable maybe be recorded several times. For example, the severity of manic/irritable episodes (data field id = 5674 [1]) was recorded at four-time points: initial assessment visit, first repeat assessment visit, imaging visit, and first repeat imaging visit). The UK Biobank terminology defines each time point as an “instance”, and the measurements recorded at a single time point as “arrays”. The column format is defined as $field\_id - instance.array$ [2]. For example, column ”5674-0.0” refers to the initial assessment visit, while ”5674-1.0” refers to the first subsequent visit. Data field id = 5076 (”Number of letters correct in round (right)” [3]) presents an example where multiple measurements were recorded during a single visit e.g. ”5076-0.0”, ”5076-0.1”, ”5076-0.2”.

Variables types include continuous, integer, categorical (one possible answer), categorical (multiple possible answers), text, time, and compound[4]. Here compound describes a variable type where a set of variables are required to describe some compound property, for example, an applanation curve (describes the reflected light signal from the cornea, field id = 5266). A categorical (one possible answer) field could refer to a binary response (e.g., yes or no), but could also refer to multiple responses from which a participant could only select one. The categorical (multiple possible answers) fields allowed participants to provide multiple answers, for example, the response option to the question ”Which of the following do you never eat?” included ”eggs”, ”dairy products”, ”wheat products”, ”sugar”, ”I eat all of the above”, or ”Prefer not to answer”.

##### 1.1.3 Data engineering strategy

We developed a de novo data engineering pipeline, and subsequently performed feature selection using an elastic net algorithm, where identified features were replicated in a 20% independent hold-out set using a permuted change in c-statistics to estimate feature importance. This feature selection pipeline was independently applied to six outcomes: coronary heart disease (CHD), ischemic stroke, heart failure (HF), atrial fibrillation (AF), CVD (combining CHD, stroke, and peripheral arterial disease (PAD)), and a broader definition of CVD+HF+AF, including HF and AF - outcomes which are more common in people with diabetes [5], [6].

The raw UKB data was curated using a purpose-built data-engineering pipeline returning quality-controlled tabular data allowing for subsequent analysis; see Appendix Figure 1. Briefly, the data engineering procedure was split into two parts: 1) basic filtering and standardization: excluding meta-data, non-baseline features, and curating retained fields (e.g., combining information recorded in separate fields, see Appendix Methods, Appendix Figure 2), and 2) type-specific transformation: creating binary variables from categorical fields, removing invariant variables and categories with too few occurrences (Appendix Methods, Appendix Figure 3).

In the *initial filtering and standardization* (Appendix Figure 2) fields were excluded if they did not present data measured at enrolment. Additionally, pilot fields, representing a subset of the data available in the non-pilot data, and categorical fields with one possible answer, but with multiple data items measured at the same time were removed. Subsequently, data fields containing meta-data, such as time (representing time of measurement), text (representing information such as device type or id) were removed from the analysis. Given the often complex inter-relationship between compound fields, these were excluded from the analysis. Furthermore, country-specific variables not recorded for all considered countries (England, Scotland, Wales) were removed. The mean value was used for continuous (floating point numbers e.g. participant height) and integer (whole number e.g. participant age) fields with multiple measurements at the same assessment visit. Additionally, related data fields also including data fields available for all considered countries were merged; see Appendix Table 3 and Appendix Table 4. For example, a single combine rule links all data fields that measure the participant's weight (data field ids: 3160 (manual entry) from body size measures, 23098 from body composition by impedance) or glucose (data field ids: 30740 from blood chemistry, 23470 from NMR spectroscopy). Next, 98 semantic rules (consisting of conditioning and affected data fields) were manually determined and reviewed (by KD and AFS) to mitigate the issue of missing responses wherever it was possible; see Appendix Table 5. This is due to the nature of the UKB, where not all of the participants were asked the same set of questions, the follow-up question may depend on a participant's previous answer. The semantic rules were defined using a related field to determine a missing value. For example, data field id = 1249 ("Past tobacco smoking") was collected from participants except those who indicated they currently smoke on most or all days, as defined by their answers to data field = 1239 ("Current tobacco smoking"). Finally, procedural responses such as "Do not know" or "Prefer not to answer" were replaced by missing values (Appendix Table 6).

The initial filtering step was followed by a *type-specific transformation* step (Appendix Figure 3 adopted from [7]), where retained data was further processed. For example, continuous data were screened for a percentage of constant values, where fields where more than 20% of participants had the same value, were additionally scrutinised to determine whether this information might be better represented as binary or categorical coding, or removed entirely. For questions where participants could provide multiple answers the categorical variable was converted to a set of binary indicator variables. For example, for the variable considering reasons of reduced smoking (data field = 6158 [8]), with values ”illness or ill health”, “doctor's advice”, “health precaution”, or ”financial reasons”, I created binary variables e.g. ”illness or ill health”: $\{True, False\}$, or “doctor's advice”: $\{True, False\}$, and so on.

While the UKB was designed to capture information about all participants, irrespective of possible clinical diagnoses, some of the data is affected by missing data. Furthermore, some of the data fields are only available (by design) for a subset of participants. This issue was partially addressed by the data engineering steps, where a part of the missing values was filled using semantic rules (see Appendix Table 5). Additionally, features that were retained after the data engineering step were screened for the percentage of missingness, excluding categories (i.e., containing multiple variables) where over 40% of the values were missing; see Appendix Tables 2, 7. The dataset was enhanced by basic demographic variables such as age, sex, and country of origin information.

##### 1.1.4 Statistical analysis

In the current analysis we exclusively focussed on measurements which were offered to all participants, with the limited number of missing data points imputed using the R package MICE [9].

After randomly splitting the data into 80% for training, and 20% for testing, the training data was used to prune features which showed a very weak univariable association with the considered outcomes. Specifically, for each of the 6 CVD outcomes we calculated the Spearman's correlation and dropped variables with a p-value equal or larger than 0.80. Subsequently, we identified variables with an absolute pairwise Spearman's correlation of 0.70 or larger – indicative of multicollinearity, dropping one of the variables that made up a multicollinear pair.

##### 1.1.5 Identification of mutually independent features

To identify novel CVD-related features we leveraged a generalized linear model with a binomial distribution and an elastic net penalty (reflecting a combination of L1 and L2 penalty), removing features not associating with the considered outcome. Elastic net models will automatically perform a feature selection step, based on the required regularization necessary to optimize the c-statistic, where for the same number of candidate features but a smaller number of cases the expected regularization will increase (and hence the number of selected features will decrease) [10]. Ten-fold cross-validation, stratified by case (people who developed CVD) and control status (people who did not develop CVD), was used to optimize model hyper-parameters - e.g., to decide on the amount and type (the L1/L2 ratio) of penalization, using the available training data.

##### 1.1.6 Feature importance

The feature importance of each selected variable was evaluated using a permutation feature importance algorithm (using 10 permutations), quantifying the change in the c-statistic. Features were subsequently ranked by their c-statistic change, stratified by outcome type (CVD+AF+HF, CVD, CHD, HF, AF, Isch. Stroke) and a participant group (”wo T2DM/CVD”, ”w T2DM”, ”w T2DM&CVD”). To provide an aggregate evaluation of the importance of each feature, we additionally recorded how often a feature was selected in each of the 6 available outcomes and calculated the summed c-statistic per participant group. The feature importance permutation algorithm was applied to the test data to provide independent replication, and as additional assurance against over-fitting. Here we dropped features with a zero or negative feature importance in the test data, indicating a failure to replicate. As age and sex are well-known and dominant CVD risk factors, the main text focussed on the remaining features, noting these remaining features are conditionally independent of age and sex. Performance of age and sex was included along with a complete list of the identified features in the relevant Appendix Tables 8 – 10 and Appendix Data 1.

Next, we identified the rank of features used in any the following three clinically used prediction models: ASCVD [11], QRISK3 [12] and the Framingham 1998 [13] score. Here we focussed on predicting 10-years risk of CVD, differentiated between whether a variable included in these three models was selected by our feature selection pipeline, and if so what its rank across the three participant groups.

To illustrate the benefit of these novel risk factors for risk classification we calculated the net-reclassification index [14] for the onset of CVD or HF, comparing predictions based on the classical risk factors employed by the ASCVD [11], QRISK3 [12] and the Framingham 1998 [13] models, compared to consideration of both novel and classical risk factors. For these calculations we applied risk cut-offs of below 10% risk, between 10 and 20%, and above 20%, and focussed on the wo T2DM/CVD and w T2DM participant subgroups given that the participants of w T2DM/CVD predominantly received predicted risk above 20%. We calculated the number of cases which appropriately received a higher risk by calculating the difference between the proportion of cases receiving a higher risk and the proportion of cases receiving a lower risk, multiplying the difference by a 1000.

Despite this manuscript not necessarily focussing on deriving a novel prediction model, the models were evaluated in terms of their discriminative (c-statistic) ability using the independent test dataset. The differences in discriminate performance between training and testing were used as a metric of model over-fit, where an overfitted model reflects sample size-specific peculiarities, instead of generalizable findings. This complemented the feature importance estimates calculated on the independent test dataset, which by using the independent test set, were unaffected by any potential model overfit.

#### 1.2 Results

##### 1.2.1 CVD features selected in all three participants groups

To show the commonalities between all three groups, we next identified features that were selected for all three participant subgroups, stratifying by CVD outcome type; Appendix Data 2. The number of common features ranged from 14 for CHD to 44 for AF. For CHD the common features recorded information on familial disease history (e.g., familial history of heart disease), self-reported general health (e.g. quit smoking due to illness, insomnia, narcolepsy), socio-economic factors (educational attainment), and biochemistry (e.g., HDL-C, urinary microalbumin). For AF common features included anthropometrics (e.g. estimated trunk mass, height), self-reported health (e.g. recent illness or injury), familial disease history (sibling with Parkinson's disease), biochemistry (e.g. urate, albumin, total bilirubin, neutrophil percentage, cystatin C. The HF common features covered socioeconomic factors (e.g., on disability allowance, "housing: renting/owning", unable to work due to illness), biochemistry (e.g. urate, Cystatin C, HbA1c, RBW), self-reported health (e.g. health satisfaction) and familial disease history (e.g. paternal history of dementia, recent illness or injury). For Ischemic stroke the common features included systolic blood pressure, biochemistry (e.g. HbA1c, Cystatin C, testosterone), diet (intake of raw vegetables, intake of cheese, not eating

##### 1.2.2 Difference in discriminative performance between training and testing samples

The elastic net models were derived in a training and testing split, where performance in the test set provides an unbiased estimate of performance, unaffected by potential model over-fitting which would also impact feature selection. To identify potential problematic analyses, affected by over-fitting, we calculated the difference in c-statistic between the training and testing sets, where a large difference is indicative of model over-fit. The difference was smaller than 0.1 for all analyses, aside from a relatively large difference of 0.18 for the model predicting Ischaemic stroke in people with T2DM and CVD at enrolment, which correlated with a small number of cases (316) relative to the number of candidate predictors (238); see Appendix Table 11, Appendix Table 16.

#### 1.3 Tables

**Table 1:** Outcome definitions based on the CALIBER dataset.

| **Type of cardiovascular disease** | **Link to the CALIBER research platform** |
| --- | --- |
| Fatal and non-fatal MI | https://www.caliberresearch.org/portal/show/phenotype_mi<br>https://www.caliberresearch.org/portal/show/phenotype_chd_nos |
| Fatal or non-fatal Stroke | https://www.caliberresearch.org/portal/show/phenotype_stroke_intracerebral_haem<br>https://www.caliberresearch.org/portal/show/phenotype_stroke_ischaemic<br>https://www.caliberresearch.org/portal/show/phenotype_stroke_nos<br>https://www.caliberresearch.org/portal/show/phenotype_stroke_subarachnoid |
| Fatal peripheral vascular disease | https://www.caliberresearch.org/portal/show/phenotype_pad |
| Sudden Cardiac Death | https://www.caliberresearch.org/portal/show/phenotype_scd |
| HF | https://www.caliberresearch.org/portal/show/phenotype_hf |
| AF | https://www.caliberresearch.org/portal/show/phenotype_af |
| Ischemic stroke | https://www.caliberresearch.org/portal/show/phenotype_stroke_ischaemic,<br>https://www.caliberresearch.org/portal/show/phenotype_stroke_nos |
| Haemorrhagic stroke | https://www.caliberresearch.org/portal/show/phenotype_stroke_intracerebral_haem,<br>https://www.caliberresearch.org/portal/show/phenotype_stroke_subarachnoid |

**Table 2:** UK Biobank category identifiers included in a study.

| Category Id | Category description |
|---|---|
| 100009 | Body composition by impedance - Anthropometry - Physical measures - Assessment Centre |
| 100010 | Body size measures - Anthropometry - Physical measures - Assessment Centre |
| 100014 | Autorefraction - Eye measures - Physical measures - Assessment Centre |
| 100015 | Intraocular pressure - Eye measures - Physical measures - Assessment Centre |
| 100017 | Visual acuity - Eye measures - Physical measures - Assessment Centre |
| 100019 | Hand grip strength - Physical measures - Assessment Centre |
| 100027 | Fluid intelligence / reasoning - Cognitive function - Assessment Centre |
| 100028 | Lights pattern memory - Cognitive function - Assessment Centre |
| 100030 | Pairs matching - Cognitive function - Assessment Centre |
| 100031 | Prospective memory - Cognitive function - Assessment Centre |
| 100032 | Reaction time - Cognitive function - Assessment Centre |
| 100034 | Family history - Touchscreen - Assessment Centre |
| 100052 | Diet - Lifestyle and environment - Touchscreen - Assessment Centre |
| 100053 | Electronic device use - Lifestyle and environment - Touchscreen - Assessment Centre |
| 100057 | Sleep - Lifestyle and environment - Touchscreen - Assessment Centre |
| 100058 | Smoking - Lifestyle and environment - Touchscreen - Assessment Centre |
| 100060 | Mental health - Psychosocial factors - Touchscreen - Assessment Centre |
| 100063 | Education - Sociodemographics - Touchscreen - Assessment Centre |
| 100064 | Employment - Sociodemographics - Touchscreen - Assessment Centre |
| 100065 | Ethnicity - Sociodemographics - Touchscreen - Assessment Centre |
| 100066 | Household - Sociodemographics - Touchscreen - Assessment Centre |
| 100067 | Other sociodemographic factors - Sociodemographics - Touchscreen - Assessment Centre |
| 100077 | Word production - Cognitive function - Assessment Centre |
| 100081 | Blood count - Blood assays - Biological samples |
| 100083 | Urine assays - Biological samples |
| 100099 | Eye surgery/complications - Eye measures - Physical measures - Assessment Centre |
| 114 | Residential air pollution - Local environment - Additional exposures |
| 17518 | Blood biochemistry - Blood assays - Biological samples |
| 54 | MET Scores - Physical activity - Lifestyle and environment - Touchscreen - Assessment Centre |
| 76 | Indices of Multiple Deprivation - Baseline characteristics - Population characteristics |
| 100011 | Assessment centre - Physical measures - Blood pressure |

**Table 3:** List of the UK Biobank data fields that were combined.

| Data field identifiers | Unified data field name |
|---|---|
| 26414,26431,26421 | Education score |
| 26412,26429,26419 | Employment score |
| 26413,26430,26420 | Health score |
| 26415,26432,26423 | Housing score |
| 26411,26428,26418 | Income score |
| 26410,26427,26426 | Index of Multiple Deprivation |
| 3160,23098,21002 | Weight |
| 21001,23104 | Body mass index (BMI) |
| 4079,94 | Diastolic blood pressure |
| 95,102 | Pulse rate |
| 4080,93 | Systolic blood pressure |

**Table 4:** Identifiers of country-specific UK Biobank data fields

| Data field ids | Data field description |
|---|---|
| 26414 (England), 26431 (Scotland), 26421 (Wales) | Education score |
| 26412 (England), 26429 (Scotland), 26419 (Wales) | Employment score |
| 26413 (England), 26430 (Scotland), 26420 (Wales) | Health score |
| 26415 (England), 26432 (Scotland), 26423 (Wales) | Housing score |
| 26411 (England), 26428 (Scotland), 26418 (Wales) | Income score |
| 26410 (England), 26427 (Scotland), 26426 (Wales) | Index of Multiple Deprivation |

**Table 5:** Semantic rules to process and standardize UK Biobank data fields

| Conditioning data field | Conditioning triggering values | Affected data field | Baseline value for affected data field |
|---|---|---|---|
| Type of accommodation lived in (670) | (4 : Sheltered accommodation)—(5 : Care home) | How are people in household related to participant (6141) | (8 : Other unrelated) |
| Type of accommodation lived in (670) | (4 : Sheltered accommodation)—(5 : Care home) | Average total household income before tax (738) | Not applicable |
| Type of accommodation lived in (670) | (4 : Sheltered accommodation)—(5 : Care home) | Number in household: for sheler or care home? (709) | Not applicable |
| Type of accommodation lived in (670) | (4 : Sheltered accommodation)—(5 : Care home) | Number of vehicles in household (728) | 1 |
| Type of accommodation lived in (670) | (4 : Sheltered accommodation)—(5 : Care home) | Own or rent accommodation lived in (680) | (3 : Rent - from local authority, local council, housing association) |
| Type of accommodation lived in (670) | (4 : Sheltered accommodation)—(5 : Care home) | Number in household (709) | Not applicable |
| Current employment status (6142) | (3 : Looking after home and/or family) | Distance between home and job workplace (796) | 0 |
| Current employment status (6142) | (2 : Retired)—(4 : Unable to work because of sickness or disability)—(5 : Unemployed)—(6 : Doing unpaid or voluntary work)—(7 : Full or part-time student)—(-7 : None of the above) | Distance between home and job workplace (796) | Not applicable |
| Current employment status (6142) | (2 : Retired)—(3 : Looking after home and/or family)—(4 : Unable to work because of sickness or disability)—(5 : Unemployed)—(6 : Doing unpaid or voluntary work)—(7 : Full or part-time student)—(-7 : None of the above) | NaN | (1 : Never/rarely) |
| Current employment status (6142) | (2 : Retired)—(3 : Looking after home and/or family)—(4 : Unable to work because of sickness or disability)—(5 : Unemployed)—(6 : Doing unpaid or voluntary work)—(7 : Full or part-time student)—(-7 : None of the above) | Transport type for commuting to job workplace (6143) | Not applicable |
| Current employment status (6142) | (2 : Retired)—(3 : Looking after home and/or family)—(4 : Unable to work because of sickness or disability)—(5 : Unemployed)—(6 : Doing unpaid or voluntary work)—(7 : Full or part-time student)—(-7 : None of the above) | Frequency of travelling from home to job workplace (777) | (0 : work from home) |
| Current employment status (6142) | (2 : Retired)—(3 : Looking after home and/or family)—(4 : Unable to work because of sickness or disability)—(5 : Unemployed)—(6 : Doing unpaid or voluntary work)—(7 : Full or part-time student)—(-7 : None of the above) | Job involves heavy manual or physical work (816) | (1 : Never/rarely) |
| Current employment status (6142) | (2 : Retired)—(3 : Looking after home and/or family)—(4 : Unable to work because of sickness or disability)—(5 : Unemployed)—(6 : Doing unpaid or voluntary work)—(7 : Full or part-time student)—(-7 : None of the above) | Job involves mainly walking or standing (806) | (1 : Never/rarely) |
| Current employment status (6142) | (2 : Retired)—(3 : Looking after home and/or family)—(4 : Unable to work because of sickness or disability)—(5 : Unemployed)—(6 : Doing unpaid or voluntary work)—(7 : Full or part-time student)—(-7 : None of the above) | Job involves shift work (826) | (1 : Never/rarely) |
| Current employment status (6142) | (2 : Retired)—(4 : Unable to work because of sickness or disability)—(5 : Unemployed)—(6 : Doing unpaid or voluntary work)—(7 : Full or part-time student)—(-7 : None of the above) | Length of working week for main job (767) | 0 |
| Current employment status (6142) | (3 : Looking after home and/or family) | Length of working week for main job (767) | 168 |

Continued on next page

**Table 5 – continued from previous page**

| Conditioning data field | Conditioning triggering values | Affected data field | Baseline value for affected data field |
|---|---|---|---|
| Current employment status (6142) | (2 : Retired)—(3 : Looking after home and/or family)—(4 : Unable to work because of sickness or disability)—(5 : Unemployed)—(6 : Doing unpaid or voluntary work)—(7 : Full or part-time student)—(-7 : None of the above) | Time employed in main current job (757) | Not applicable |
| Current tobacco smoking (1239) | (2 : Only occasionally)—(0 : No) | Difficulty not smoking for 1 day (3476) | (1 : Very easy) |
| Current tobacco smoking (1239) | (2 : Only occasionally)—(0 : No) | Number of cigarettes currently smoked daily (current cigarette smokers) (3456) | 0 |
| Current tobacco smoking (1239) | (2 : Only occasionally)—(0 : No) | Previously smoked cigarettes on most/all days (5959) | Not applicable |
| Current tobacco smoking (1239) | (2 : Only occasionally)—(0 : No) | Time from waking to first cigarette (3466) | Not applicable |
| Current tobacco smoking (1239) | (2 : Only occasionally)—(0 : No) | Why reduced smoking (6158) | Not applicable |
| Current tobacco smoking (1239) | (2 : Only occasionally)—(0 : No) | Age started smoking in current smokers (3436) | Not applicable |
| Current tobacco smoking (1239) | (2 : Only occasionally)—(0 : No) | Ever tried to stop smoking (3486) | Not applicable |
| Current tobacco smoking (1239) | (1 : Yes, on most or all days) | Exposure to tobacco smoke at home (1269) | (168 : hours/week) |
| Current tobacco smoking (1239) | (1 : Yes, on most or all days) | Exposure to tobacco smoke outside home (1279) | (168 : hours/week) |
| Current tobacco smoking (1239) | (1 : Yes, on most or all days) | Past tobacco smoking (1249) | (1 : Smoked on all or most days) |
| Current tobacco smoking (1239) | (2 : Only occasionally)—(0 : No) | Smoking compared to 10 years previous (3506) | Not applicable |
| Current tobacco smoking (1239) | (1 : Yes, on most or all days) | Smoking/smokers in household (1259) | Not applicable |
| Current tobacco smoking (1239) | (2 : Only occasionally)—(0 : No) | Type of tobacco currently smoked (3446) | Not applicable |
| Current tobacco smoking (1239) | (2 : Only occasionally)—(0 : No) | Wants to stop smoking (3496) | Not applicable |
| Processed meat intake (1349) | (1 : Less than once a week)—(2 : Once a week)—(3 : 2-4 times a week)—(4 : 5-6 times a week)—(5 : Once or more daily) | Age when last ate meat (3680) | current_age |
| Poultry intake (1359) | (1 : Less than once a week)—(2 : Once a week)—(3 : 2-4 times a week)—(4 : 5-6 times a week)—(5 : Once or more daily) | Age when last ate meat (3680) | current_age |
| Beef intake (1369) | (1 : Less than once a week)—(2 : Once a week)—(3 : 2-4 times a week)—(4 : 5-6 times a week)—(5 : Once or more daily) | Age when last ate meat (3680) | current_age |
| Lamb/mutton intake (1379) | (1 : Less than once a week)—(2 : Once a week)—(3 : 2-4 times a week)—(4 : 5-6 times a week)—(5 : Once or more daily) | Age when last ate meat (3680) | current_age |
| Pork intake (1389) | (1 : Less than once a week)—(2 : Once a week)—(3 : 2-4 times a week)—(4 : 5-6 times a week)—(5 : Once or more daily) | Age when last ate meat (3680) | current_age |
| Ever manic/hyper for 2 days (4642) | (0 : No) | Length of longest manic/irritable episode (5663) | Not applicable |
| Ever manic/hyper for 2 days (4642) | (0 : No) | Manic/hyper symptoms (6156) | Not applicable |
| Ever manic/hyper for 2 days (4642) | (0 : No) | Severity of manic/irritable episodes (5674) | Not applicable |
| Ever highly irritable/argumentative for 2 days (4653) | (0 : No) | Length of longest manic/irritable episode (5663) | Not applicable |

Continued on next page

**Table 5 – continued from previous page**

| Conditioning data field | Conditioning triggering values | Affected data field | Baseline value for affected data field |
|---|---|---|---|
| Ever highly irritable/argumentative for 2 days (4653) | (0 : No) | Manic/hyper symptoms (6156) | Not applicable |
| Ever highly irritable/argumentative for 2 days (4653) | (0 : No) | Severity of manic/irritable episodes (5674) | Not applicable |
| Ever stopped smoking for 6+ months (2907) | (0 : No) | Likelihood of resuming smoking (2936) | Not applicable |
| Ever stopped smoking for 6+ months (2907) | (0 : No) | Number of unsuccessful stop-smoking attempts (2926) | Not applicable |
| Ever stopped smoking for 6+ months (2907) | (0 : No) | Why stopped smoking (6157) | Not applicable |
| Type of tobacco previously smoked (2877) | (3 : Cigars or pipes)—(-7 : None of the above) | Number of cigarettes previously smoked daily (2887) | 0 |
| Smoking compared to 10 years previous (3506) | (1 : More nowadays?)—(2 : About the same?) | Why reduced smoking (6158) | Not applicable |
| Previously smoked cigarettes on most/all days (5959) | (0 : No) | Age stopped smoking cigarettes (current cigar/pipe or previous cigarette smoker) (6194) | Not applicable |
| Previously smoked cigarettes on most/all days (5959) | (0 : No) | Number of cigarettes previously smoked daily (current cigar/pipe smokers) (6183) | Not applicable |
| Type of tobacco currently smoked (3446) | (1 : Manufactured cigarettes)—(2 : Hand-rolled cigarettes)—(-7 : None of the above) | Age stopped smoking cigarettes (current cigar/pipe or previous cigarette smoker) (6194) | Not applicable |
| Type of tobacco currently smoked (3446) | (3 : Cigars or pipes)—(-7 : None of the above) | Difficulty not smoking for 1 day (3476) | Not applicable |
| Type of tobacco currently smoked (3446) | (3 : Cigars or pipes)—(-7 : None of the above) | Number of cigarettes currently smoked daily (current cigarette smokers) (3456) | Not applicable |
| Type of tobacco currently smoked (3446) | (1 : Manufactured cigarettes)—(2 : Hand-rolled cigarettes)—(-7 : None of the above) | Number of cigarettes previously smoked daily (current cigar/pipe smokers) (6183) | 0 |
| Type of tobacco currently smoked (3446) | (1 : Manufactured cigarettes)—(2 : Hand-rolled cigarettes)—(-7 : None of the above) | Previously smoked cigarettes on most/all days (5959) | Not applicable |
| Type of tobacco currently smoked (3446) | (3 : Cigars or pipes)—(-7 : None of the above) | Time from waking to first cigarette (3466) | Not applicable |
| Past tobacco smoking (1249) | (2 : Smoked occasionally)—(3 : Just tried once or twice)—(4 : I have never smoked) | Likelihood of resuming smoking (2936) | Not applicable |
| Past tobacco smoking (1249) | (2 : Smoked occasionally)—(3 : Just tried once or twice)—(4 : I have never smoked) | Number of cigarettes previously smoked daily (2887) | 0 |
| Past tobacco smoking (1249) | (2 : Smoked occasionally)—(3 : Just tried once or twice)—(4 : I have never smoked) | Number of unsuccessful stop-smoking attempts (2926) | Not applicable |
| Past tobacco smoking (1249) | (2 : Smoked occasionally)—(3 : Just tried once or twice)—(4 : I have never smoked) | Why stopped smoking (6157) | Not applicable |
| Past tobacco smoking (1249) | (2 : Smoked occasionally)—(3 : Just tried once or twice)—(4 : I have never smoked) | Age started smoking in former smokers (2867) | Not applicable |
| Past tobacco smoking (1249) | (2 : Smoked occasionally)—(3 : Just tried once or twice)—(4 : I have never smoked) | Age stopped smoking (2897) | Not applicable |
| Past tobacco smoking (1249) | (2 : Smoked occasionally)—(3 : Just tried once or twice)—(4 : I have never smoked) | Ever stopped smoking for 6+ months (2907) | Not applicable |

Continued on next page

**Table 5 – continued from previous page**

| Conditioning data field | Conditioning triggering values | Affected data field | Baseline value for affected data field |
|---|---|---|---|
| Past tobacco smoking (1249) | (1 : Smoked on most or all days) | Light smokers, at least 100 smokes in lifetime (2644) | (1 : yes) |
| Past tobacco smoking (1249) | (4 : I have never smoked) | Light smokers, at least 100 smokes in lifetime (2644) | (0 : no) |
| Past tobacco smoking (1249) | (2 : Smoked occasionally)—(3 : Just tried once or twice)—(4 : I have never smoked) | Type of tobacco previously smoked (2877) | Not applicable |
| Adopted as a child (1767) | (0 : No) | Adopted mother still alive (3942) | Not applicable |
| Adopted mother still alive (3942) | (0 : No) | Mother's age (1845) | Not applicable |
| Adopted mother still alive (3942) | (1 : Yes) | Mother's age at death (3526) | Not applicable |
| Mother still alive (1835) | (0 : No) | Mother's age (1845) | Not applicable |
| Mother still alive (1835) | (1 : Yes) | Mother's age at death (3526) | Not applicable |
| Adopted as a child (1767) | (0 : No) | Adopted father still alive (3912) | Not applicable |
| Adopted father still alive (3912) | (0 : No) | Father's age (2946) | Not applicable |
| Adopted father still alive (3912) | (1 : Yes) | Father's age at death (1807) | Not applicable |
| Father still alive (1797) | (0 : No) | Father's age (2946) | Not applicable |
| Father still alive (1797) | (1 : Yes) | Father's age at death (1807) | Not applicable |
| Job involves shift work (826) | (1 : Never/rarely) | Job involves night shift work (3426) | (1 : Never/rarely) |
| Frequency of travelling from home to job workplace (777) | NaN | Distance between home and job workplace (796) | 0 |
| Frequency of travelling from home to job workplace (777) | NaN | Transport type for commuting to job workplace (6143) | Not applicable |
| Adopted as a child (1767) | (0 : No) | Illnesses of adopted father (20112) | Not applicable |
| Adopted as a child (1767) | (0 : No) | Illnesses of adopted mother (20113) | Not applicable |
| Adopted as a child (1767) | (0 : No) | Illnesses of adopted siblings (20114) | Not applicable |
| Bipolar disorder status (20122) | NaN | Bipolar disorder status (20122) | Not collected |
| Single episode of probable major depression (20123) | NaN | Single episode of probable major depression (20123) | Not collected |
| Probable recurrent major depression (moderate) (20124) | NaN | Probable recurrent major depression (moderate) (20124) | Not collected |
| Probable recurrent major depression (severe) (20125) | NaN | Probable recurrent major depression (severe) (20125) | Not collected |
| Bipolar and major depression status (20126) | NaN | Bipolar and major depression status (20126) | Not collected |
| Myopia diagnosis (20262) | NaN | Myopia diagnosis (20262) | Not collected |
| Country of birth (UK/elsewhere) (1647) | (6 : Elsewhere) | Year immigrated to UK (United Kingdom) (3659) | Not applicable |
| Adopted as a child (1767) | (0 : No) | Number of adopted brothers (3972) | Not applicable |

Continued on next page

**Table 5 – continued from previous page**

| Conditioning data field | Conditioning triggering values | Affected data field | Baseline value for affected data field |
|---|---|---|---|
| Adopted as a child (1767) | (0 : No) | Number of adopted sisters (3982) | Not applicable |
| Ever depressed for a whole week (4598) | (0 : No) | Longest period of depression (4609) | Not applicable |
| Ever depressed for a whole week (4598) | (0 : No) | Number of depression episodes (4620) | Not applicable |
| Adopted as a child (1767) | (1 : Yes) | Number of older siblings (5057) | Not applicable |
| Ever unenthusiastic/disinterested for a whole week (4631) | (0 : No) | Ever unenthusiastic/disinterested for a whole week (5375) | Not applicable |
| Ever unenthusiastic/disinterested for a whole week (4631) | (0 : No) | Number of unenthusiastic/disinterested episodes (5386) | Not applicable |
| Past tobacco smoking (1249) | (4 : I have never smoked) | Pack years of smoking (20161) | 0 |
| Past tobacco smoking (1249) | (4 : I have never smoked) | Pack years adult smoking as proportion of life span exposed to smoking (20162) | 0 |
| Microalbumin in urine result flag (30505) | (<6.7: ”below 6.7 mg”) | Microalbumin in urine (30500) | 0 |

**Table 6:** UK Biobank (UKB) question answers replaced by missing values.

| UKB response identifier | UBK description |
|---|---|
| -1 | Question not asked due to previous answers |
| -3 | Prefer not to answer |
| -7 | None of the above |
| -11 | Do not know (group 1) |
| -21 | Do not know (group 2) |
| -1 | Do not know |
| -1 | Participant skipped/abandonded |
| -17 | None of the above (group 1) |
| -23 | Prefer not to answer (group 2) |
| -27 | None of the above (group 2) |
| -5 | Not sure |
| -13 | Prefer not to answer (group 1) |
| -10 | Less than once a week |

**Table 7:** Average percentage of missing data for each UK Biobank categories after data engineering .

| Category Id | Category description | Average % missing |
|---|---|---|
| 100009 | Body composition by impedance - Anthropometry - Physical measures - Assessment Centre | 2.05 |
| 100010 | Body size measures - Anthropometry - Physical measures - Assessment Centre | 14.72 |
| 100011 | Assessment centre - Physical measures - Blood pressure | 34.42 |
| 100014 | Autorefraction - Eye measures - Physical measures - Assessment Centre | 80.01 |
| 100015 | Intraocular pressure - Eye measures - Physical measures - Assessment Centre | 77.70 |
| 100017 | Visual acuity - Eye measures - Physical measures - Assessment Centre | 77.31 |
| 100019 | Hand grip strength - Physical measures - Assessment Centre | 0.67 |
| 100027 | Fluid intelligence / reasoning - Cognitive function - Assessment Centre | 78.99 |
| 100030 | Pairs matching - Cognitive function - Assessment Centre | 0.93 |
| 100031 | Prospective memory - Cognitive function - Assessment Centre | 65.88 |
| 100032 | Reaction time - Cognitive function - Assessment Centre | 1.49 |
| 100034 | Family history - Touchscreen - Assessment Centre | 7.04 |
| 100052 | Diet - Lifestyle and environment - Touchscreen - Assessment Centre | 4.49 |
| 100053 | Electronic device use - Lifestyle and environment - Touchscreen - Assessment Centre | 11.50 |
| 100057 | Sleep - Lifestyle and environment - Touchscreen - Assessment Centre | 3.10 |
| 100058 | Smoking - Lifestyle and environment - Touchscreen - Assessment Centre | 10.06 |
| 100060 | Mental health - Psychosocial factors - Touchscreen - Assessment Centre | 33.86 |
| 100063 | Education - Sociodemographics - Touchscreen - Assessment Centre | 5.62 |
| 100064 | Employment - Sociodemographics - Touchscreen - Assessment Centre | 7.12 |
| 100065 | Ethnicity - Sociodemographics - Touchscreen - Assessment Centre | 0.55 |
| 100066 | Household - Sociodemographics - Touchscreen - Assessment Centre | 24.60 |
| 100067 | Other sociodemographic factors - Sociodemographics - Touchscreen - Assessment Centre | 16.75 |
| 100081 | Blood count - Blood assays - Biological samples | 5.32 |
| 100083 | Urine assays - Biological samples | 4.09 |
| 100099 | Eye surgery/complications - Eye measures - Physical measures - Assessment Centre | 90.12 |
| 114 | Residential air pollution - Local environment - Additional exposures | 4.72 |
| 17518 | Blood biochemistry - Blood assays - Biological samples | 15.79 |
| 54 | MET Scores - Physical activity - Lifestyle and environment - Touchscreen - Assessment Centre | 19.70 |
| 76 | Indices of Multiple Deprivation - Baseline characteristics - Population characteristics | 13.90 |

**Table 8:** Feature importance of top 20 variables identified by the Elastic Net model for ”wo T2DM/CVD” group.

| Data field id | Description | Mean permuted feature importance (change in c-stat) | Std after permuted feature importance (change in c-stat) | Outcome |
|---|---|---|---|---|
| age_defined_baseline | Age (years) | 0.086469 | 0.001262 | CVD+AF+HF |
| genetic_sex | Sex | 0.007480 | 0.000170 | CVD+AF+HF |
| 23130 | Estimated trunk mass | 0.005181 | 0.000176 | CVD+AF+HF |
| 4080 | SBP | 0.002723 | 0.000135 | CVD+AF+HF |
| 20107 | Father: heart disease | 0.001901 | 0.000133 | CVD+AF+HF |
| 30760 | HDL-C | 0.001062 | 0.000089 | CVD+AF+HF |
| 30720 | Cystatin C | 0.001711 | 0.000174 | CVD+AF+HF |
| 30850 | Testosterone | 0.000775 | 0.000046 | CVD+AF+HF |
| 20110 | Mother: heart disease | 0.001963 | 0.000196 | CVD+AF+HF |
| 30750 | HbA1c | 0.001567 | 0.000174 | CVD+AF+HF |
| 30500 | Urine microalbumin | 0.001028 | 0.000071 | CVD+AF+HF |
| 30070 | RDW | 0.000903 | 0.000033 | CVD+AF+HF |
| 30880 | Plasma urate | 0.000831 | 0.000110 | CVD+AF+HF |
| 20111 | Sibling: heart disease | 0.001091 | 0.000089 | CVD+AF+HF |
| 23128 | Fat mass (trunk) | 0.000783 | 0.000069 | CVD+AF+HF |
| 2080 | Self-reported: recent tiredness | 0.001216 | 0.000070 | CVD+AF+HF |
| 30140 | Neutrophill count | 0.001138 | 0.000080 | CVD+AF+HF |
| 4548 | Self-reported: health satisfaction | 0.001397 | 0.000092 | CVD+AF+HF |
| 6145 | Recent: illness—injury—assault | 0.001280 | 0.000095 | CVD+AF+HF |
| 30050 | MCH | 0.000629 | 0.000072 | CVD+AF+HF |
| age_defined_baseline | Age (years) | 0.069037 | 0.001483 | CVD |
| genetic_sex | Sex | 0.024033 | 0.000360 | CVD |
| 4080 | SBP | 0.006624 | 0.000169 | CVD |
| 20107 | Father: heart disease | 0.003187 | 0.000184 | CVD |
| 30760 | HDL-C | 0.003061 | 0.000122 | CVD |
| 30720 | Cystatin C | 0.001593 | 0.000233 | CVD |
| 20110 | Mother: heart disease | 0.002144 | 0.000222 | CVD |
| 30750 | HbA1c | 0.003057 | 0.000257 | CVD |
| 30600 | Plasma albumin | 0.000438 | 0.000066 | CVD |
| 30500 | Urine microalbumin | 0.000710 | 0.000066 | CVD |
| 30070 | RDW | 0.000448 | 0.000057 | CVD |
| 20111 | Sibling: heart disease | 0.001567 | 0.000113 | CVD |
| 2080 | Self-reported: recent tiredness | 0.001441 | 0.000133 | CVD |
| 30140 | Neutrophill count | 0.000878 | 0.000097 | CVD |
| 4548 | Self-reported: health satisfaction | 0.001465 | 0.000120 | CVD |
| 6145 | Recent: illness—injury—assault | 0.000751 | 0.000089 | CVD |
| 26413 | Health score | 0.000665 | 0.000068 | CVD |
| 6157 | Quit smoking due to illness | 0.000622 | 0.000081 | CVD |
| 30790 | LP[a] | 0.000532 | 0.000079 | CVD |
| 47 | Grip strength (right hand) | 0.000463 | 0.000053 | CVD |
| age_defined_baseline | Age (years) | 0.064730 | 0.001508 | CHD |
| genetic_sex | Sex | 0.030887 | 0.000532 | CHD |
| 4080 | SBP | 0.006163 | 0.000170 | CHD |
| 20107 | Father: heart disease | 0.005119 | 0.000195 | CHD |
| 30760 | HDL-C | 0.004174 | 0.000095 | CHD |
| 30720 | Cystatin C | 0.001101 | 0.000220 | CHD |
| 20110 | Mother: heart disease | 0.003168 | 0.000321 | CHD |
| 30750 | HbA1c | 0.003062 | 0.000267 | CHD |
| 30880 | Plasma urate | 0.000440 | 0.000132 | CHD |
| 20111 | Sibling: heart disease | 0.001970 | 0.000121 | CHD |
| 23128 | Fat mass (trunk) | 0.000475 | 0.000081 | CHD |
| 2080 | Self-reported: recent tiredness | 0.001813 | 0.000173 | CHD |
| 30140 | Neutrophill count | 0.001250 | 0.000134 | CHD |

Continued on next page

**Table 8 Continued from previous page**

| Data field id | Description | Mean permuted feature importance (change in c-stat) | Std after permuted feature importance (change in c-stat) | Outcome |
|---|---|---|---|---|
| 4548 | Self-reported: health satisfaction | 0.001642 | 0.000155 | CHD |
| 6145 | Recent: illness—injury—assault | 0.000689 | 0.000060 | CHD |
| 26413 | Health score | 0.000449 | 0.000059 | CHD |
| 6157 | Quit smoking due to illness | 0.000802 | 0.000100 | CHD |
| 30780 | LDL-C | 0.000520 | 0.000114 | CHD |
| 30790 | LP[a] | 0.000704 | 0.000114 | CHD |
| 1408 | Cheese consumption | 0.000532 | 0.000074 | CHD |
| age_defined_baseline | Age (years) | 0.081982 | 0.002885 | Is. Stroke |
| genetic_sex | Sex | 0.001664 | 0.000242 | Is. Stroke |
| 4080 | SBP | 0.006972 | 0.000543 | Is. Stroke |
| 30720 | Cystatin C | 0.003629 | 0.000363 | Is. Stroke |
| 30850 | Testosterone | 0.003510 | 0.000322 | Is. Stroke |
| 30750 | HbA1c | 0.001294 | 0.000422 | Is. Stroke |
| 30600 | Plasma albumin | 0.002748 | 0.000511 | Is. Stroke |
| 30500 | Urine microalbumin | 0.001479 | 0.000264 | Is. Stroke |
| 30070 | RDW | 0.000522 | 0.000251 | Is. Stroke |
| 2080 | Self-reported: recent tiredness | 0.000709 | 0.000186 | Is. Stroke |
| 6145 | Recent: illness—injury—assault | 0.001025 | 0.000215 | Is. Stroke |
| 26413 | Health score | 0.001359 | 0.000378 | Is. Stroke |
| 6146 | Receives: disability allowance | 0.000866 | 0.000170 | Is. Stroke |
| 3506 | Difference smoking (10-years) | 0.000835 | 0.000257 | Is. Stroke |
| 30780 | LDL-C | 0.000590 | 0.000187 | Is. Stroke |
| 30050 | MCH | 0.000592 | 0.000177 | Is. Stroke |
| 6144 | Self-reported: does not avoid particular foods | 0.000596 | 0.000241 | Is. Stroke |
| 20107 | Father: stroke | 0.000594 | 0.000151 | Is. Stroke |
| 24011 | Traffic nearest major road | 0.000540 | 0.000147 | Is. Stroke |
| 20107 | Father: bowel cancer | 0.000526 | 0.000177 | Is. Stroke |
| age_defined_baseline | Age (years) | 0.109217 | 0.001754 | AF |
| 23130 | Estimated trunk mass | 0.024359 | 0.000583 | AF |
| 20107 | Father: heart disease | 0.000703 | 0.000074 | AF |
| 30850 | Testosterone | 0.001367 | 0.000085 | AF |
| 20110 | Mother: heart disease | 0.001420 | 0.000197 | AF |
| 30500 | Urine microalbumin | 0.001539 | 0.000092 | AF |
| 30070 | RDW | 0.001359 | 0.000125 | AF |
| 30880 | Plasma urate | 0.002172 | 0.000189 | AF |
| 23128 | Fat mass (trunk) | 0.001859 | 0.000127 | AF |
| 30140 | Neutrophill count | 0.001752 | 0.000164 | AF |
| 4548 | Self-reported: health satisfaction | 0.000885 | 0.000075 | AF |
| 6145 | Recent: illness—injury—assault | 0.001637 | 0.000098 | AF |
| 30830 | SHBG | 0.001619 | 0.000176 | AF |
| 30100 | MPV | 0.001316 | 0.000141 | AF |
| 51 | Seated height | 0.001222 | 0.000138 | AF |
| 30530 | Urine sodium | 0.001152 | 0.000124 | AF |
| 30090 | PCT | 0.000887 | 0.000094 | AF |
| 30670 | Plasma Urea | 0.000832 | 0.000113 | AF |
| 30840 | Bilirubin | 0.000785 | 0.000137 | AF |
| 30780 | LDL-C | 0.000743 | 0.000088 | AF |
| age_defined_baseline | Age (years) | 0.068911 | 0.002678 | HF |
| genetic_sex | Sex | 0.003644 | 0.000533 | HF |
| 23130 | Estimated trunk mass | 0.005822 | 0.000484 | HF |
| 4080 | SBP | 0.003287 | 0.000513 | HF |
| 30720 | Cystatin C | 0.003409 | 0.000534 | HF |
| 30750 | HbA1c | 0.001476 | 0.000216 | HF |
| 30600 | Plasma albumin | 0.001585 | 0.000122 | HF |

Continued on next page

**Table 8 Continued from previous page**

| Data field id | Description | Mean permuted feature importance (change in c-stat) | Std after permuted feature importance (change in c-stat) | Outcome |
|---|---|---|---|---|
| 30500 | Urine microalbumin | 0.002440 | 0.000283 | HF |
| 30070 | RDW | 0.002354 | 0.000273 | HF |
| 30880 | Plasma urate | 0.001803 | 0.000332 | HF |
| 6146 | Disability parking permit (blue badge) | 0.001892 | 0.000239 | HF |
| 30140 | Neutrophill count | 0.000657 | 0.000196 | HF |
| 4548 | Self-reported: health satisfaction | 0.001265 | 0.000255 | HF |
| 6145 | Recent: illness—injury—assault | 0.000685 | 0.000124 | HF |
| 30830 | SHBG | 0.000871 | 0.000110 | HF |
| 26413 | Health score | 0.000915 | 0.000164 | HF |
| 30270 | MSCV | 0.000996 | 0.000203 | HF |
| 6157 | Quit smoking due to illness | 0.000655 | 0.000135 | HF |
| 30840 | Bilirubin | 0.000697 | 0.000192 | HF |
| 30200 | Neutrophill (%) | 0.000777 | 0.000166 | HF |

n.b. The permutation feature importance assesses the c-statistic change in the test data; iteratively the values of each variables were randomly assigned to an individual after which the c-statistic was re-estimated with these permuted data and the difference in performance used as an estimate of variable contribution to the model's predictive potential.

**Table 9:** Feature importance of top 20 variables identified by the Elastic Net model for "w T2DM" group.

| Data field id | Description | Mean permuted feature importance (change in c-stat) | Std after permuted feature importance (change in c-stat) | Outcome |
|---|---|---|---|---|
| age_defined_baseline | Age (years) | 0.040588 | 0.004557 | CVD+AF+HF |
| 30720 | Cystatin C | 0.020745 | 0.002039 | CVD+AF+HF |
| genetic_sex | Sex | 0.010660 | 0.001410 | CVD+AF+HF |
| 4548 | Self-reported: health satisfaction | 0.006048 | 0.001321 | CVD+AF+HF |
| 30600 | Plasma albumin | 0.008787 | 0.001654 | CVD+AF+HF |
| 4080 | SBP | 0.001444 | 0.000388 | CVD+AF+HF |
| 20107 | Father: heart disease | 0.000577 | 0.000186 | CVD+AF+HF |
| 1200 | Self-reported: insomnia | 0.001367 | 0.000266 | CVD+AF+HF |
| 20110 | Mother: chronic bronchitis—emphysema | 0.000667 | 0.000233 | CVD+AF+HF |
| 20110 | Mother: heart disease | 0.002714 | 0.000578 | CVD+AF+HF |
| 6146 | Disability parking permit (blue badge) | 0.003020 | 0.001000 | CVD+AF+HF |
| 23130 | Estimated trunk mass | 0.002217 | 0.001037 | CVD+AF+HF |
| 6145 | Recent: illness—injury—assault | 0.002210 | 0.000561 | CVD+AF+HF |
| 796 | Distance home to workplace | 0.000820 | 0.000388 | CVD+AF+HF |
| 26413 | Health score | 0.000694 | 0.000250 | CVD+AF+HF |
| 4079 | DBP | 0.000710 | 0.000240 | CVD+AF+HF |
| 6142 | Unemployed due to illness/disability | 0.001275 | 0.000393 | CVD+AF+HF |
| 6138 | A(S) levels qualifications | 0.000669 | 0.000130 | CVD+AF+HF |
| 6157 | Quit smoking due to illness | 0.001237 | 0.000215 | CVD+AF+HF |
| 30770 | IGF-1 | 0.000498 | 0.000339 | CVD+AF+HF |
| age_defined_baseline | Age (years) | 0.026725 | 0.005273 | CVD |
| 30720 | Cystatin C | 0.019278 | 0.002801 | CVD |
| genetic_sex | Sex | 0.019466 | 0.002635 | CVD |
| 4548 | Self-reported: health satisfaction | 0.009867 | 0.001797 | CVD |
| 30750 | HbA1c | 0.001579 | 0.000249 | CVD |
| 30600 | Plasma albumin | 0.005292 | 0.001321 | CVD |
| 4080 | SBP | 0.001807 | 0.000420 | CVD |
| 20107 | Father: heart disease | 0.000578 | 0.000192 | CVD |
| 1200 | Self-reported: insomnia | 0.002765 | 0.000694 | CVD |
| 20110 | Mother: chronic bronchitis—emphysema | 0.000498 | 0.000202 | CVD |
| 20110 | Mother: heart disease | 0.001937 | 0.000616 | CVD |
| 6146 | Disability parking permit (blue badge) | 0.003033 | 0.001061 | CVD |
| 6145 | Recent: illness—injury—assault | 0.002375 | 0.000746 | CVD |
| 26413 | Health score | 0.001282 | 0.000377 | CVD |
| 6142 | Unemployed due to illness/disability | 0.001519 | 0.000576 | CVD |
| 6138 | A(S) levels qualifications | 0.001377 | 0.000278 | CVD |
| 6157 | Quit smoking due to illness | 0.001360 | 0.000434 | CVD |
| 1408 | Cheese consumption | 0.001346 | 0.000260 | CVD |
| 30500 | Urine microalbumin | 0.000526 | 0.000567 | CVD |
| 30140 | Neutrophill count | 0.000661 | 0.000374 | CVD |
| age_defined_baseline | Age (years) | 0.017316 | 0.004318 | CHD |
| 30720 | Cystatin C | 0.019868 | 0.003271 | CHD |
| genetic_sex | Sex | 0.016927 | 0.002343 | CHD |
| 4548 | Self-reported: health satisfaction | 0.014177 | 0.002608 | CHD |
| 30600 | Plasma albumin | 0.002314 | 0.000608 | CHD |
| 20107 | Father: heart disease | 0.000914 | 0.000285 | CHD |
| 1200 | Self-reported: insomnia | 0.005303 | 0.001229 | CHD |
| 2080 | Self-reported: recent tiredness | 0.000962 | 0.000340 | CHD |
| 20110 | Mother: heart disease | 0.000940 | 0.000298 | CHD |
| 6146 | Disability parking permit (blue badge) | 0.001755 | 0.001041 | CHD |
| 6145 | Recent: illness—injury—assault | 0.001823 | 0.000938 | CHD |
| 796 | Distance home to workplace | 0.002305 | 0.000753 | CHD |
| 26413 | Health score | 0.001213 | 0.000293 | CHD |

Continued on next page

**Table 9 Continued from previous page**

| Data field id | Description | Mean permuted feature importance (change in c-stat) | Std after permuted feature importance (change in c-stat) | Outcome |
|---|---|---|---|---|
| 6138 | A(S) levels qualifications | 0.001283 | 0.000220 | CHD |
| 6157 | Quit smoking due to illness | 0.000922 | 0.000432 | CHD |
| 30500 | Urine microalbumin | 0.001282 | 0.000631 | CHD |
| 2010 | Suffers from nerves | 0.000884 | 0.000268 | CHD |
| 30760 | HDL-C | 0.000862 | 0.000361 | CHD |
| 1220 | Self-reported: narcolepsy | 0.000478 | 0.000235 | CHD |
| 404 | Reaction time: duration to press button | 0.000449 | 0.000233 | CHD |
| age_defined_baseline | Age (years) | 0.048557 | 0.005868 | Is. Stroke |
| 30850 | Testosterone | 0.021875 | 0.004335 | Is. Stroke |
| 30720 | Cystatin C | 0.010568 | 0.002869 | Is. Stroke |
| 30750 | HbA1c | 0.011310 | 0.002513 | Is. Stroke |
| 30600 | Plasma albumin | 0.005678 | 0.002529 | Is. Stroke |
| 30860 | Total plasma protein | 0.008407 | 0.002913 | Is. Stroke |
| 6157 | Quite smoking due to financial reasons | 0.007803 | 0.003727 | Is. Stroke |
| 20107 | Father: heart disease | 0.006596 | 0.002116 | Is. Stroke |
| 1299 | Raw vegetable consumption | 0.006442 | 0.001663 | Is. Stroke |
| 20110 | Mother: chronic bronchitis—emphysema | 0.004334 | 0.001756 | Is. Stroke |
| 30300 | HLR | 0.004112 | 0.001786 | Is. Stroke |
| 6146 | Receives: disability allowance | 0.003978 | 0.001189 | Is. Stroke |
| 30680 | Plasma calcium | 0.003971 | 0.001201 | Is. Stroke |
| 2080 | Self-reported: recent tiredness | 0.003834 | 0.001471 | Is. Stroke |
| 30530 | Urine sodium | 0.002228 | 0.000490 | Is. Stroke |
| 30830 | SHBG | 0.003312 | 0.001458 | Is. Stroke |
| 20111 | Sibling: stroke | 0.002718 | 0.001111 | Is. Stroke |
| 6138 | O-levels/GCSEs | 0.002256 | 0.000691 | Is. Stroke |
| 26413 | Health score | 0.002234 | 0.001132 | Is. Stroke |
| 6145 | Recent: death of partner | 0.002174 | 0.001796 | Is. Stroke |
| age_defined_baseline | Age (years) | 0.036644 | 0.004067 | AF |
| 30720 | Cystatin C | 0.003214 | 0.001083 | AF |
| genetic_sex | Sex | 0.001556 | 0.001097 | AF |
| 30880 | Plasma urate | 0.003633 | 0.001067 | AF |
| 30600 | Plasma albumin | 0.009726 | 0.002140 | AF |
| 6157 | Quite smoking due to financial reasons | 0.001219 | 0.000238 | AF |
| 4080 | SBP | 0.002218 | 0.001074 | AF |
| 23128 | Fat mass (trunk) | 0.004055 | 0.001535 | AF |
| 30200 | Neutrophill (%) | 0.003812 | 0.000897 | AF |
| 30530 | Urine sodium | 0.001992 | 0.000407 | AF |
| 30070 | RDW | 0.001334 | 0.000649 | AF |
| 20110 | Mother: heart disease | 0.003165 | 0.000627 | AF |
| 6142 | Looking after home and/or family | 0.002568 | 0.000591 | AF |
| 23130 | Estimated trunk mass | 0.002394 | 0.001669 | AF |
| 4079 | DBP | 0.001570 | 0.000504 | AF |
| 20111 | Sibling: breast cancer | 0.001524 | 0.000438 | AF |
| 20110 | Mother: stroke | 0.001467 | 0.000421 | AF |
| 20111 | Sibling: severe depression | 0.001437 | 0.000590 | AF |
| 24013 | Residential traffic load | 0.001237 | 0.000116 | AF |
| 1528 | No. glasses of water | 0.001169 | 0.000367 | AF |
| age_defined_baseline | Age (years) | 0.037528 | 0.006198 | HF |
| 30720 | Cystatin C | 0.015868 | 0.003386 | HF |
| genetic_sex | Sex | 0.020708 | 0.003584 | HF |
| 30880 | Plasma urate | 0.011659 | 0.002783 | HF |
| 30750 | HbA1c | 0.011228 | 0.002473 | HF |
| 30600 | Plasma albumin | 0.005252 | 0.001809 | HF |
| 4080 | SBP | 0.006644 | 0.001599 | HF |
| 23128 | Fat mass (trunk) | 0.006583 | 0.002337 | HF |

Continued on next page

**Table 9 Continued from previous page**

| Data field id | Description | Mean permuted feature importance (change in c-stat) | Std after permuted feature importance (change in c-stat) | Outcome |
|---|---|---|---|---|
| 20107 | Father: lung cancer | 0.004030 | 0.000668 | HF |
| 30530 | Urine sodium | 0.003644 | 0.001237 | HF |
| 1140 | Change in mobile phone use | 0.003573 | 0.001187 | HF |
| 30070 | RDW | 0.003493 | 0.001026 | HF |
| 20111 | Sibling: prostate cancer | 0.003379 | 0.001379 | HF |
| 22036 | Recommended level of physical activity | 0.003311 | 0.001284 | HF |
| 6144 | Self-reported: does not avoid particular foods | 0.003234 | 0.001631 | HF |
| 47 | Grip strength (right hand) | 0.003214 | 0.001303 | HF |
| 6144 | Does not consume wheat | 0.002861 | 0.001218 | HF |
| 6139 | Heating: gas hob—gas cooker | 0.002646 | 0.000912 | HF |
| 30790 | LP[a] | 0.002461 | 0.001087 | HF |
| 680 | Housing: renting/owning | 0.001816 | 0.000539 | HF |

n.b. The permutation feature importance assesses the c-statistic change in the test data; iteratively the values of each variables were randomly assigned to an individual after which the c-statistic was re-estimated with these permuted data and the difference in performance used as an estimate of variable contribution to the model's predictive potential.

**Table 10:** Feature importance of top 20 variables identified by the Elastic Net model for "w T2DM&CVD" group.

| Data field id | Description | Mean permuted feature importance (change in c-stat) | Std after permuted feature importance (change in c-stat) | Outcome |
|---|---|---|---|---|
| age_defined_baseline | Age (years) | 0.029004 | 0.004052 | CVD+AF+HF |
| genetic_sex | Sex | 0.011679 | 0.001641 | CVD+AF+HF |
| 30070 | RDW | 0.002037 | 0.001087 | CVD+AF+HF |
| 6157 | Quit smoking due to illness | 0.011211 | 0.003668 | CVD+AF+HF |
| 4079 | DBP | 0.006832 | 0.002462 | CVD+AF+HF |
| 6146 | Disability parking permit (blue badge) | 0.007786 | 0.002327 | CVD+AF+HF |
| 30030 | Haematocrit (%) | 0.004988 | 0.001380 | CVD+AF+HF |
| 4537 | Work/job satisfaction | 0.005176 | 0.001820 | CVD+AF+HF |
| 30520 | Urine potassium | 0.005159 | 0.001644 | CVD+AF+HF |
| 20107 | Father: chronic bronchitis/emphysema | 0.003467 | 0.001964 | CVD+AF+HF |
| 30840 | Bilirubin | 0.001939 | 0.001126 | CVD+AF+HF |
| 1970 | Self-reported: nervous person | 0.003465 | 0.001889 | CVD+AF+HF |
| 30500 | Urine microalbumin | 0.001911 | 0.001355 | CVD+AF+HF |
| England | Origin country: England | 0.003388 | 0.001148 | CVD+AF+HF |
| 30200 | Neutrophill (%) | 0.003541 | 0.001040 | CVD+AF+HF |
| 20111 | Sibling: heart disease | 0.002519 | 0.001895 | CVD+AF+HF |
| 2080 | Self-reported: recent tiredness | 0.001979 | 0.001156 | CVD+AF+HF |
| 1359 | Poultry consumption | 0.002029 | 0.000920 | CVD+AF+HF |
| 1408 | Cheese consumption | 0.001970 | 0.000504 | CVD+AF+HF |
| 1220 | Self-reported: narcolepsy | 0.001763 | 0.001023 | CVD+AF+HF |
| age_defined_baseline | Age (years) | 0.014740 | 0.002776 | CVD |
| genetic_sex | Sex | 0.008375 | 0.001945 | CVD |
| 6157 | Quit smoking due to illness | 0.003395 | 0.001978 | CVD |
| 4079 | DBP | 0.010578 | 0.002424 | CVD |
| 6146 | Disability parking permit (blue badge) | 0.002358 | 0.001522 | CVD |
| 30030 | Haematocrit (%) | 0.007448 | 0.001833 | CVD |
| 4537 | Work/job satisfaction | 0.002467 | 0.000876 | CVD |
| 30520 | Urine potassium | 0.003375 | 0.001098 | CVD |
| 20107 | Father: chronic bronchitis/emphysema | 0.004532 | 0.001571 | CVD |
| 1160 | Sleep (hours) | 0.002494 | 0.000816 | CVD |
| 1970 | Self-reported: nervous person | 0.004055 | 0.001623 | CVD |
| 1309 | Fruit consumption | 0.003127 | 0.001273 | CVD |
| 30500 | Urine microalbumin | 0.001969 | 0.001095 | CVD |
| 20110 | Mother: heart disease | 0.002756 | 0.000851 | CVD |
| 95 | Pulse rate | 0.002034 | 0.000514 | CVD |
| England | Origin country: England | 0.003593 | 0.000876 | CVD |
| 1920 | Self-reported: suffers from mood swings | 0.002740 | 0.000863 | CVD |
| 20111 | Sibling: heart disease | 0.002665 | 0.002118 | CVD |
| 6138 | CSE-equivalent | 0.002112 | 0.001061 | CVD |
| 1548 | Dietary variation | 0.001645 | 0.000756 | CVD |
| age_defined_baseline | Age (years) | 0.017597 | 0.003140 | CHD |
| genetic_sex | Sex | 0.020849 | 0.002662 | CHD |
| 6157 | Quit smoking due to illness | 0.004098 | 0.001596 | CHD |
| 4079 | DBP | 0.011007 | 0.002755 | CHD |
| 6146 | Disability parking permit (blue badge) | 0.002521 | 0.001127 | CHD |
| 30030 | Haematocrit (%) | 0.006538 | 0.001825 | CHD |
| 400 | Pairs matching: exercise time | 0.003486 | 0.001009 | CHD |
| 30520 | Urine potassium | 0.004627 | 0.001173 | CHD |
| 20107 | Father: heart disease | 0.004570 | 0.001951 | CHD |
| 20107 | Father: chronic bronchitis/emphysema | 0.004529 | 0.001635 | CHD |
| 1160 | Sleep (hours) | 0.004321 | 0.001122 | CHD |
| 1970 | Self-reported: nervous person | 0.002508 | 0.001179 | CHD |
| 1309 | Fruit consumption | 0.004041 | 0.001275 | CHD |

Continued on next page

**Table 10 Continued from previous page**

| Data field id | Description | Mean permuted feature importance (change in c-stat) | Std after permuted feature importance (change in c-stat) | Outcome |
|---|---|---|---|---|
| 30500 | Urine microalbumin | 0.003982 | 0.001733 | CHD |
| 20110 | Mother: heart disease | 0.003709 | 0.001048 | CHD |
| 95 | Pulse rate | 0.003692 | 0.001060 | CHD |
| England | Origin country: England | 0.003459 | 0.000871 | CHD |
| 1920 | Self-reported: suffers from mood swings | 0.002216 | 0.000694 | CHD |
| 6138 | A(S) levels qualifications | 0.002341 | 0.001090 | CHD |
| 1478 | Salt intake | 0.002186 | 0.001264 | CHD |
| age_defined_baseline | Age (years) | 0.007408 | 0.001214 | Is. Stroke |
| 30070 | RDW | 0.007840 | 0.001776 | Is. Stroke |
| 6143 | Commute: walking | 0.010064 | 0.001367 | Is. Stroke |
| 6143 | Commute: public transport | 0.009787 | 0.003650 | Is. Stroke |
| 398 | Pairs matching: correct matches | 0.009755 | 0.002575 | Is. Stroke |
| 4080 | SBP | 0.007802 | 0.002176 | Is. Stroke |
| 30810 | Phosphate | 0.007449 | 0.002507 | Is. Stroke |
| 26414 | Education score | 0.006923 | 0.001747 | Is. Stroke |
| 30730 | GGT | 0.006654 | 0.002567 | Is. Stroke |
| 1873 | No. brothers | 0.006533 | 0.003009 | Is. Stroke |
| Scotland | Origin country: Scotland | 0.006219 | 0.001421 | Is. Stroke |
| 47 | Grip strength (right hand) | 0.005743 | 0.001445 | Is. Stroke |
| 845 | Age completed education | 0.005572 | 0.001970 | Is. Stroke |
| 400 | Pairs matching: exercise time | 0.005276 | 0.002028 | Is. Stroke |
| 1528 | No. glasses of water | 0.005250 | 0.002167 | Is. Stroke |
| 30710 | CRP | 0.004918 | 0.001786 | Is. Stroke |
| 30740 | Glucose | 0.004902 | 0.001587 | Is. Stroke |
| 2010 | Suffers from nerves | 0.004798 | 0.001827 | Is. Stroke |
| 30780 | LDL-C | 0.004687 | 0.001716 | Is. Stroke |
| 26413 | Health score | 0.004639 | 0.001341 | Is. Stroke |
| age_defined_baseline | Age (years) | 0.032322 | 0.003092 | AF |
| 30070 | RDW | 0.018608 | 0.002259 | AF |
| 30880 | Plasma urate | 0.009819 | 0.002041 | AF |
| 1980 | Self-reported: worrier/anxious feelings | 0.007337 | 0.001430 | AF |
| 6158 | Smoked less due to illness | 0.005805 | 0.002548 | AF |
| 6145 | Recent: illness—injury—assault | 0.005715 | 0.001538 | AF |
| 26413 | Health score | 0.002651 | 0.001132 | AF |
| 30840 | Bilirubin | 0.004414 | 0.002216 | AF |
| 6146 | Receives: disability allowance | 0.003774 | 0.001335 | AF |
| 2080 | Self-reported: recent tiredness | 0.002337 | 0.000945 | AF |
| 6141 | Living with sibling | 0.002287 | 0.001446 | AF |
| 30680 | Plasma calcium | 0.002141 | 0.000619 | AF |
| 6138 | NVQ/HND/HNC-equivalent | 0.002126 | 0.000723 | AF |
| 23130 | Estimated trunk mass | 0.001926 | 0.001226 | AF |
| 1279 | Tobacco exposure outside home | 0.001778 | 0.001112 | AF |
| 6144 | Does not consume wheat | 0.001775 | 0.000619 | AF |
| 20023 | Mean time to correctly identify matches | 0.001749 | 0.000599 | AF |
| 30100 | MPV | 0.001692 | 0.000518 | AF |
| 30230 | NRBC | 0.001661 | 0.000785 | AF |
| 1210 | Self-reported: snoring | 0.001543 | 0.000974 | AF |
| age_defined_baseline | Age (years) | 0.011201 | 0.002871 | HF |
| 30070 | RDW | 0.008492 | 0.002170 | HF |
| 30880 | Plasma urate | 0.004547 | 0.001617 | HF |
| 6146 | Disability parking permit (blue badge) | 0.003821 | 0.001658 | HF |
| 6145 | Recent: illness—injury—assault | 0.002412 | 0.000979 | HF |
| 30750 | HbA1c | 0.005302 | 0.001059 | HF |
| 30520 | Urine potassium | 0.002758 | 0.001591 | HF |
| 20111 | Sibling: severe depression | 0.003823 | 0.001413 | HF |

Continued on next page

**Table 10 Continued from previous page**

| Data field id | Description | Mean permuted feature importance (change in c-stat) | Std after permuted feature importance (change in c-stat) | Outcome |
|---|---|---|---|---|
| 6146 | Receives: disability allowance | 0.002029 | 0.001345 | HF |
| 30200 | Neutrophill (%) | 0.002441 | 0.001401 | HF |
| 30720 | Cystatin C | 0.003353 | 0.001808 | HF |
| 30090 | PCT | 0.002940 | 0.001189 | HF |
| 30130 | Monocyte count | 0.002473 | 0.000747 | HF |
| 1478 | Salt intake | 0.001706 | 0.000709 | HF |
| 6138 | NVQ/HND/HNC-equivalent | 0.001640 | 0.000564 | HF |
| 23130 | Estimated trunk mass | 0.001919 | 0.001323 | HF |
| 1498 | Coffee consumption | 0.001857 | 0.000974 | HF |
| 1140 | Change in mobile phone use | 0.001800 | 0.000816 | HF |
| 6158 | Smoked less as a health precaution | 0.001719 | 0.000827 | HF |
| 24015 | Nearby major roads | 0.001510 | 0.000362 | HF |

n.b. The permutation feature importance assesses the c-statistic change in the test data; iteratively the values of each variables were randomly assigned to an individual after which the c-statistic was re-estimated with these permuted data and the difference in performance used as an estimate of variable contribution to the model's predictive potential.

**Table 11:** The number of variables used for training a multivariable elastic net algorithm and the number of CVD events during a 10-year follow-up period stratified by training and testing samples.

| Group | Outcome | Sample size | No. events train (%) | No. events test (%) | No. variables |
|---|---|---|---|---|---|
| wo T2DM/CVD | CVD+AF+HF | 459142 | 32306 (8.8) | 8044 (8.8) | 252 |
| w T2DM | CVD+AF+HF | 14610 | 2130 (18.2) | 541 (18.5) | 246 |
| w T2DM&CVD | CVD+AF+HF | 4432 | 2745 (77.4) | 708 (79.8) | 236 |
| wo T2DM/CVD | CVD | 459142 | 22021 (6.0) | 5447 (5.9) | 255 |
| w T2DM | CVD | 14610 | 1619 (13.9) | 415 (14.2) | 239 |
| w T2DM&CVD | CVD | 4432 | 2617 (73.8) | 669 (75.4) | 234 |
| wo T2DM/CVD | CHD | 459142 | 17394 (4.7) | 4318 (4.7) | 255 |
| w T2DM | CHD | 14610 | 1271 (10.9) | 312 (10.7) | 233 |
| w T2DM&CVD | CHD | 4432 | 2464 (69.5) | 639 (72.0) | 228 |
| wo T2DM/CVD | HF | 459142 | 3985 (1.1) | 969 (1.1) | 251 |
| w T2DM | HF | 14610 | 405 (3.5) | 91 (3.1) | 224 |
| w T2DM&CVD | HF | 4432 | 728 (20.5) | 214 (24.1) | 237 |
| wo T2DM/CVD | AF | 459142 | 15058 (4.1) | 3745 (4.1) | 257 |
| w T2DM | AF | 14610 | 915 (7.8) | 221 (7.6) | 235 |
| w T2DM&CVD | AF | 4432 | 879 (24.8) | 234 (26.4) | 237 |
| wo T2DM/CVD | Is. Stroke | 459142 | 3588 (1.0) | 841 (0.9) | 251 |
| w T2DM | Is. Stroke | 14610 | 266 (2.3) | 74 (2.5) | 228 |
| w T2DM&CVD | Is. Stroke | 4432 | 316 (8.9) | 72 (8.1) | 238 |

n.b. The UK Biobank participants were grouped based on T2DM and CVD histories at the time of enrolment: participants without a history of CVD and T2DM (wo T2DM/CVD), participants with type 2 diabetes (w T2DM), participants with a history of CVD before a T2DM diagnosis (w T2DM&CVD).

**Table 12:** Number of data fields excluded based data-driven filtering steps.

| Group | Outcome | Insufficient outcome correlation | Multicolinearity |
|---|---|---|---|
| wo T2DM/CVD | CVD+AF+HF | 9 | 121 |
| w T2DM | CVD+AF+HF | 19 | 117 |
| w T2DM&CVD | CVD+AF+HF | 33 | 112 |
| wo T2DM/CVD | CVD | 5 | 122 |
| w T2DM | CVD | 28 | 115 |
| w T2DM&CVD | CVD | 42 | 105 |
| wo T2DM/CVD | CHD | 7 | 120 |
| w T2DM | CHD | 34 | 115 |
| w T2DM&CVD | CHD | 35 | 118 |
| wo T2DM/CVD | HF | 13 | 118 |
| w T2DM | HF | 46 | 111 |
| w T2DM&CVD | HF | 36 | 108 |
| wo T2DM/CVD | AF | 8 | 117 |
| w T2DM | AF | 36 | 111 |
| w T2DM&CVD | AF | 37 | 107 |
| wo T2DM/CVD | Isch. Stroke | 17 | 114 |
| w T2DM | Isch. Stroke | 43 | 111 |
| w T2DM&CVD | Isch. Stroke | 36 | 107 |

**Table 13:** Ranks of the Qrisk3, ASCVD, and Framingham variables identified in ”wo T2DM/CVD” group for CVD outcome.

| Risk score variable | UK Biobank variable | Mean permuted feature importance (change in c-stat) | Std after permuted feature importance (change in c-stat) | Feature rank | Feature rank (%) | Qrisk3 variable | ASCVD variable | Framingham variable |
|---|---|---|---|---|---|---|---|---|
| age | age_defined_baseline | 0.069037 | 0.001483 | 1.0 | 0.53 | ✓ | ✓ | ✓ |
| sex | genetic_sex | 0.024033 | 0.00036 | 2.0 | 1.06 | ✓ | ✓ | ✓ |
| SBP | 4080-0.0 | 0.006624 | 0.000169 | 3.0 | 1.59 | ✓ | ✓ | ✓ |
| Family history of CVD (paternal) | 20107.1 | 0.003187 | 0.000184 | 4.0 | 2.12 | ✓ | | |
| HDL cholesterol | 30760-0.0 | 0.003061 | 0.000122 | 5.0 | 2.65 | ✓ | ✓ | ✓ |
| Family history of CVD (maternal) | 20110.1 | 0.002144 | 0.000222 | 7.0 | 3.7 | ✓ | | |
| Family history of CVD (sibling) | 20111.1 | 0.001567 | 0.000113 | 9.0 | 4.76 | ✓ | | |
| LDL cholesterol | 30780-0.0 | 0.000102 | 0.000041 | 57.0 | 30.16 | | ✓ | |
| DBP | 4079-0.0 | 0.000035 | 0.000022 | 102.0 | 53.97 | ✓ | ✓ | ✓ |
| Ethnicity | 21000-0.0 | 0.000023 | 0.000014 | 122.0 | 64.55 | ✓ | ✓ | |
| BMI | 21001-0.0 | not selected | | | | ✓ | | |
| Severe mental illness (severe depression) | 20125-0.0 | not selected | | | | ✓ | | |
| Towsend social deprivation score | 26410-0.0 | not selected | | | | ✓ | | |
| Severe mental illness (moderate depression) | 20123-0.0 | not selected | | | | ✓ | | |
| Total cholesterol | 30690-0.0 | not selected | | | | ✓ | ✓ | ✓ |
| Severe mental illness (moderate depression) | 20124-0.0 | not selected | | | | ✓ | | |
| Severe mental illness (biopolar disorder) | 20122-0.0 | not selected | | | | ✓ | | |
| smoking status | 20116-0.0 or 1239-0.0 | not selected | | | | ✓ | ✓ | ✓ |

n.b. Feature importance was calculated using a permuted feature importance algorithm recording the change in c-statistic. The features were ranked on their relevance for CVD prediction in ”wo T2DM/CVD” separately considering positive feature importance. Abbreviations: people without diabetes or a history of CVD at enrolment (”wo T2DM/CVD”), feature importance mean (Mean), feature importance standard deviation (Std), systolic blood pressure (SBP), cardiovascular disease (CVD), high-density lipoprotein cholesterol (HDL cholesterol), diastolic blood pressure (DBP).

**Table 14:** Ranks of the Qrisk3, ASCVD, and Framingham variables identified in ”w T2DM” group for CVD outcome.

| Risk score variable | UK Biobank variable | Mean permuted feature importance (change in c-stat) | Std after permuted feature importance (change in c-stat) | Feature rank | Feature rank (%) | Qrisk3 variable | ASCVD variable | Framingham variable |
|---|---|---|---|---|---|---|---|---|
| age | age_defined_baseline | 0.026725 | 0.005273 | 1.0 | 2.44 | ✓ | ✓ | ✓ |
| sex | genetic_sex | 0.019466 | 0.002635 | 2.0 | 4.88 | ✓ | ✓ | ✓ |
| Family history of CVD (maternal) | 20110.1 | 0.001937 | 0.000616 | 10.0 | 24.39 | ✓ | | |
| SBP | 4080-0.0 | 0.001807 | 0.00042 | 11.0 | 26.83 | ✓ | ✓ | ✓ |
| Family history of CVD (paternal) | 20107.1 | 0.000578 | 0.000192 | 19.0 | 46.34 | ✓ | | |
| DBP | 4079-0.0 | 0.000179 | 0.00007 | 30.0 | 73.17 | ✓ | ✓ | ✓ |
| Family history of CVD (sibling) | 20111.1 | 0.000112 | 0.000131 | 36.0 | 87.8 | ✓ | | |
| HDL cholesterol | 30760-0.0 | 0.000039 | 0.000043 | 40.0 | 97.56 | ✓ | ✓ | ✓ |
| BMI | 21001-0.0 | not selected | | | | ✓ | | |
| Severe mental illness (severe depression) | 20125-0.0 | not selected | | | | ✓ | | |
| Towsend social deprivation score | 26410-0.0 | not selected | | | | ✓ | | |
| Severe mental illness (moderate depression) | 20123-0.0 | not selected | | | | ✓ | | |
| LDL cholesterol | 30780-0.0 | not selected | | | | | ✓ | |
| Total cholesterol | 30690-0.0 | not selected | | | | ✓ | ✓ | ✓ |
| Ethnicity | 21000-0.0 | not selected | | | | ✓ | ✓ | |
| Severe mental illness (moderate depression) | 20124-0.0 | not selected | | | | ✓ | | |
| Severe mental illness (biopolar disorder) | 20122-0.0 | not selected | | | | ✓ | | |
| smoking status | 20116-0.0 or 1239-0.0 | not selected | | | | ✓ | ✓ | ✓ |

n.b. Feature importance was calculated using a permuted feature importance algorithm recording the change in c-statistic. The features were ranked on their relevance for CVD prediction in ”w T2DM” separately considering positive feature importance. Abbreviations: people with diabetes but without a history of CVD at enrolment (”w T2DM”), feature importance mean (Mean), feature importance standard deviation (Std), systolic blood pressure (SBP), cardiovascular disease (CVD), high-density lipoprotein cholesterol (HDL cholesterol), diastolic blood pressure (DBP).

**Table 15:** Ranks of the Qrisk3, ASCVD, and Framingham variables identified in ”w T2DM&CVD” group for CVD outcome.

| Risk score variable | UK Biobank variable | Mean permuted feature importance (change in c-stat) | Std after permuted feature importance (change in c-stat) | Feature rank | Feature rank (%) | Qrisk3 variable | ASCVD variable | Framingham variable |
|---|---|---|---|---|---|---|---|---|
| age | age_defined_baseline | 0.01474 | 0.002776 | 1.0 | 0.93 | ✓ | ✓ | ✓ |
| DBP | 4079-0.0 | 0.010578 | 0.002424 | 2.0 | 1.87 | ✓ | ✓ | ✓ |
| sex | genetic_sex | 0.008375 | 0.001945 | 3.0 | 2.8 | ✓ | ✓ | ✓ |
| Family history of CVD (maternal) | 20110.1 | 0.002756 | 0.000851 | 11.0 | 10.28 | ✓ | | |
| Family history of CVD (sibling) | 20111.1 | 0.002665 | 0.002118 | 13.0 | 12.15 | ✓ | | |
| Family history of CVD (paternal) | 20107.1 | 0.000696 | 0.0006 | 51.0 | 47.66 | ✓ | | |
| HDL cholesterol | 30760-0.0 | 0.000241 | 0.000922 | 74.0 | 69.16 | ✓ | ✓ | ✓ |
| SBP | 4080-0.0 | 0.00018 | 0.000216 | 83.0 | 77.57 | ✓ | ✓ | ✓ |
| BMI | 21001-0.0 | not selected | | | | ✓ | | |
| Severe mental illness (severe depression) | 20125-0.0 | not selected | | | | ✓ | | |
| Towsend social deprivation score | 26410-0.0 | not selected | | | | ✓ | | |
| Severe mental illness (moderate depression) | 20123-0.0 | not selected | | | | ✓ | | |
| LDL cholesterol | 30780-0.0 | not selected | | | | | ✓ | |
| Total cholesterol | 30690-0.0 | not selected | | | | ✓ | ✓ | ✓ |
| Ethnicity | 21000-0.0 | not selected | | | | ✓ | ✓ | |
| Severe mental illness (moderate depression) | 20124-0.0 | not selected | | | | ✓ | | |
| Severe mental illness (biopolar disorder) | 20122-0.0 | not selected | | | | ✓ | | |
| smoking status | 20116-0.0 or 1239-0.0 | not selected | | | | ✓ | ✓ | ✓ |

n.b. Feature importance was calculated using a permuted feature importance algorithm recording the change in c-statistic. The features were ranked on their relevance for CVD prediction in ”w T2DM&CVD” separately considering positive feature importance. Abbreviations: people with diabetes and a history of CVD at enrolment (”w T2DM&CVD”), feature importance mean (Mean), feature importance standard deviation (Std), systolic blood pressure (SBP), cardiovascular disease (CVD), high-density lipoprotein cholesterol (HDL cholesterol), diastolic blood pressure (DBP).

**Table 16:** Discrimination multivariable elastic net models predicting 10-years risk of six types of CVD.

| Group | Outcome | Train c-statistic | Test c-statistic | Diff c-statistic |
|---|---|---|---|---|
| wo T2DM/CVD | CVD+AF+HF | 0.753 (0.753; 0.753) | 0.750 (0.750; 0.750) | 0.003 |
| w T2DM | CVD+AF+HF | 0.711 (0.710; 0.711) | 0.695 (0.694; 0.696) | 0.016 |
| w T2DM&CVD | CVD+AF+HF | 0.718 (0.717; 0.718) | 0.677 (0.675; 0.678) | 0.041 |
| wo T2DM/CVD | CVD | 0.755 (0.755; 0.755) | 0.752 (0.752; 0.752) | 0.003 |
| w T2DM | CVD | 0.700 (0.700; 0.701) | 0.685 (0.684; 0.686) | 0.015 |
| w T2DM&CVD | CVD | 0.710 (0.710; 0.711) | 0.671 (0.670; 0.673) | 0.039 |
| wo T2DM/CVD | CHD | 0.757 (0.757; 0.757) | 0.753 (0.752; 0.753) | 0.004 |
| w T2DM | CHD | 0.687 (0.687; 0.688) | 0.656 (0.655; 0.657) | 0.031 |
| w T2DM&CVD | CHD | 0.701 (0.700; 0.701) | 0.659 (0.658; 0.660) | 0.042 |
| wo T2DM/CVD | HF | 0.801 (0.801; 0.802) | 0.796 (0.795; 0.797) | 0.005 |
| w T2DM | HF | 0.803 (0.802; 0.804) | 0.752 (0.750; 0.754) | 0.051 |
| w T2DM&CVD | HF | 0.766 (0.765; 0.767) | 0.746 (0.744; 0.747) | 0.020 |
| wo T2DM/CVD | AF | 0.773 (0.773; 0.773) | 0.766 (0.766; 0.766) | 0.007 |
| w T2DM | AF | 0.763 (0.762; 0.763) | 0.709 (0.707; 0.710) | 0.054 |
| w T2DM&CVD | AF | 0.731 (0.731; 0.732) | 0.689 (0.688; 0.691) | 0.042 |
| wo T2DM/CVD | Is. Stroke | 0.765 (0.765; 0.765) | 0.755 (0.754; 0.756) | 0.010 |
| w T2DM | Is. Stroke | 0.768 (0.767; 0.769) | 0.709 (0.706; 0.711) | 0.059 |
| w T2DM&CVD | Is. Stroke | 0.810 (0.809; 0.811) | 0.626 (0.624; 0.628) | 0.184 |

n.b. Individuals are stratified as followed ”wo T2DM/CVD”: participants without T2DM or CVD at baseline, “w T2DM”: participants with diabetes at baseline, “w T2DM&CVD”: participants with T2DM at baseline and a history of CVD. The analysed outcomes include cardiovascular disease including heart failure (HF) and/or atrial fibrillation (AF) (CVD+), cardiovascular disease (CVD), coronary heart disease (CHD), HF, AF, and Ischaemic Stroke. Train discrimination (c-statistic) is based on 80% of train set of the total data used for this study, while test discrimination is calculated using remaining 20% of the total dataset. Point estimates are presented alongside 95% CI.

#### 1.4 Figures

**Figure 1:** Overview of the study design pipeline.

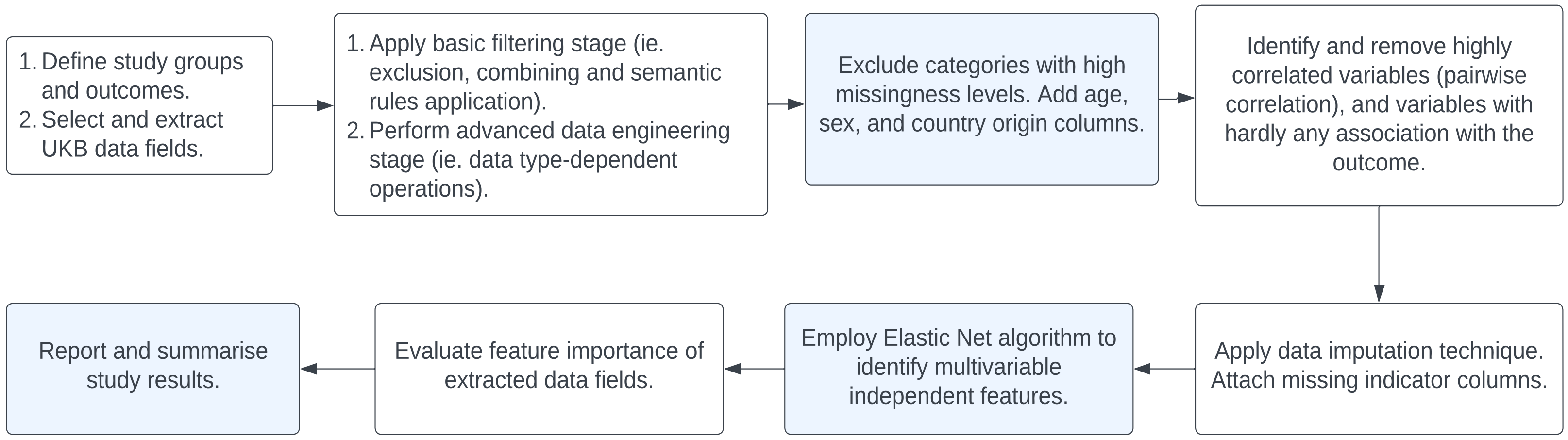

Figure 2: Flow diagram of the initial filtering and standardization step.

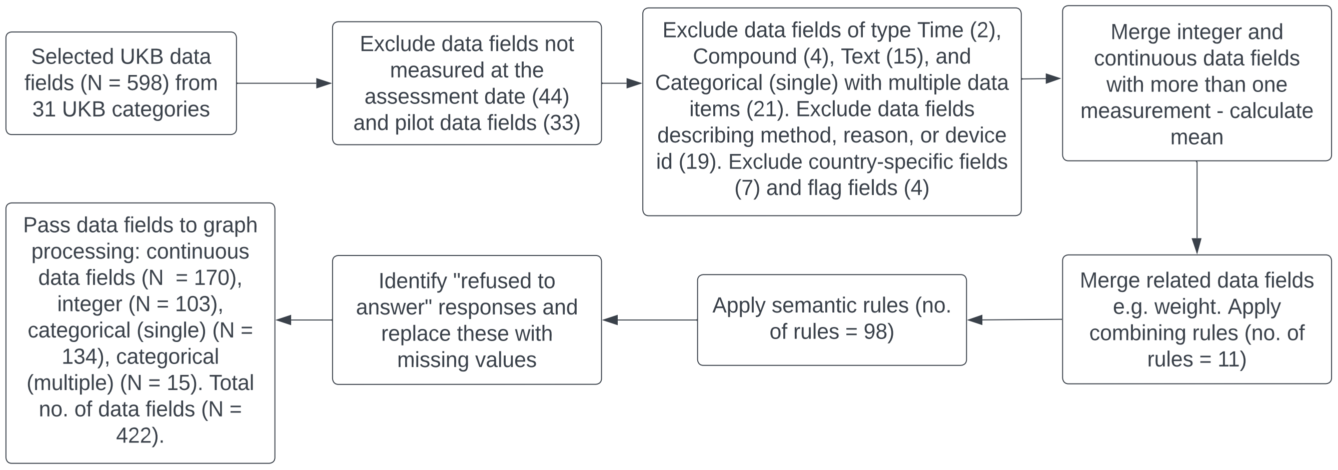

**Figure 3:** Flow diagram of data type-specific transformation step.

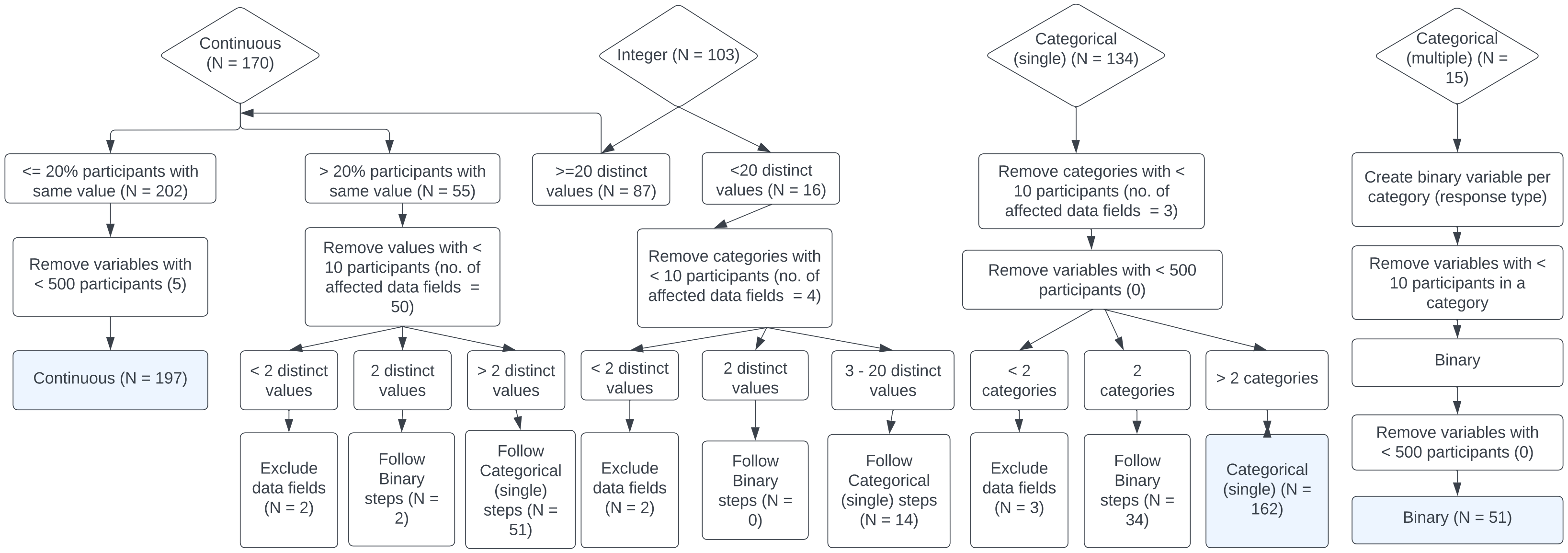

**Figure 4:** The contribution of the top features to the prediction of six facets of CVD for ”wo T2DM/CVD” group.

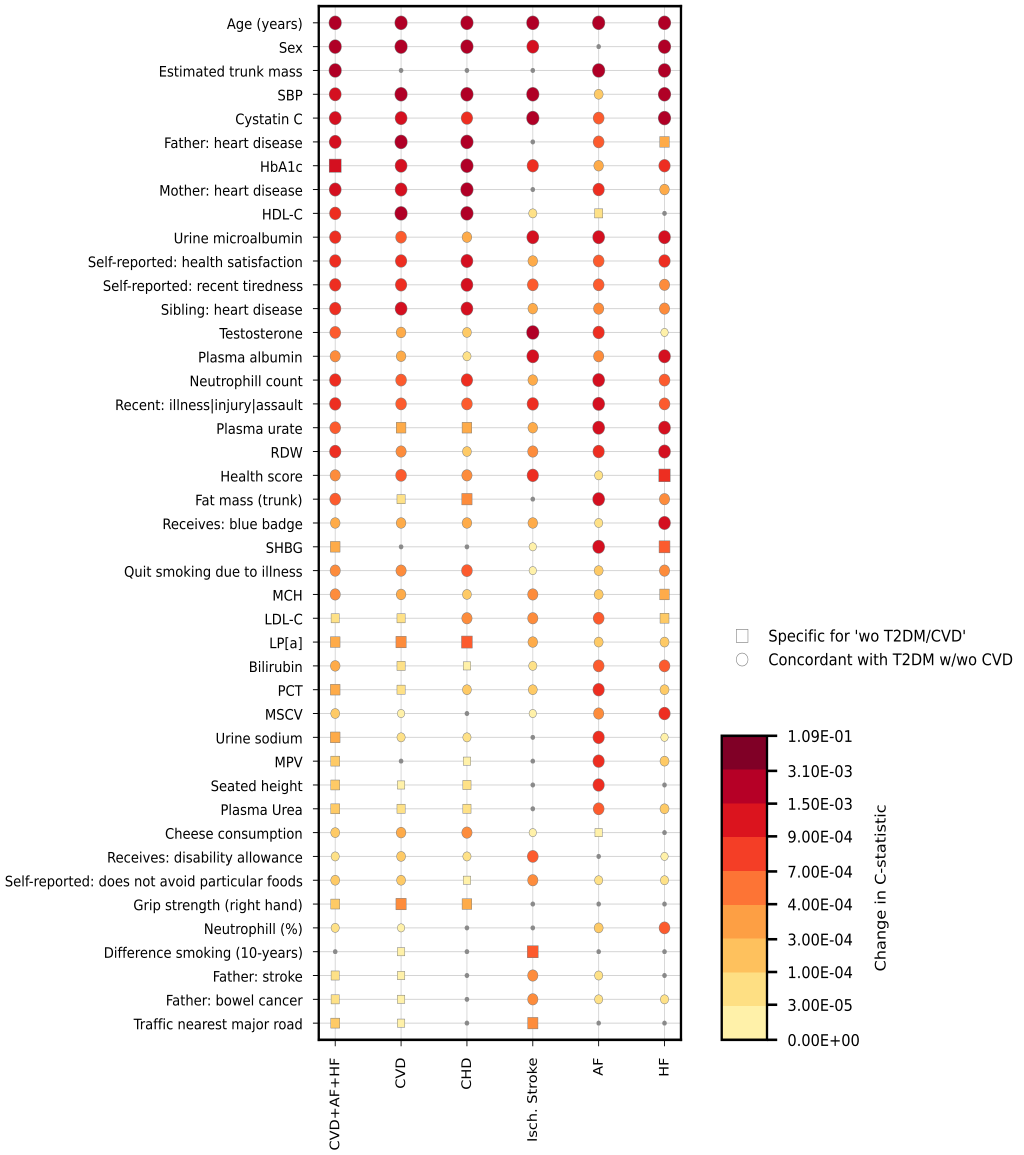

N. B. The y-axis presents the union of the top 20 features based on the c-statistic change for each of the six CVD outcomes, features in violet originate from the NMR UKB category. Plot markers (e.g. triangle, square) highlight the differences in identified features between groups. The permuted feature importance reflects the c-statistic change based on the test data; iteratively the values of each variable were randomly assigned to an individual after which the c-statistic was re-estimated with these permuted data and the difference in performance was used as an estimate of a variable contribution to the model’s predictive potential. Abbreviations: glycated haemoglobin (HbA1c), high-density lipoprotein cholesterol (HDL-C), red blood cell distribution width (RDW), systolic blood pressure (SBP), sex hormone-binding globulin (SHBG), mean corpuscular haemoglobin (MCH), lipoprotein A (LP[a]), low-density lipoprotein cholesterol (LDL-C), platelet crit (PCT), mean platelet (thrombocyte) volume (MPV), mean sphered cell volume (MSCV).

**Figure 5:** The contribution of the top 60 features to the prediction of six facets of CVD for "w T2DM" group.

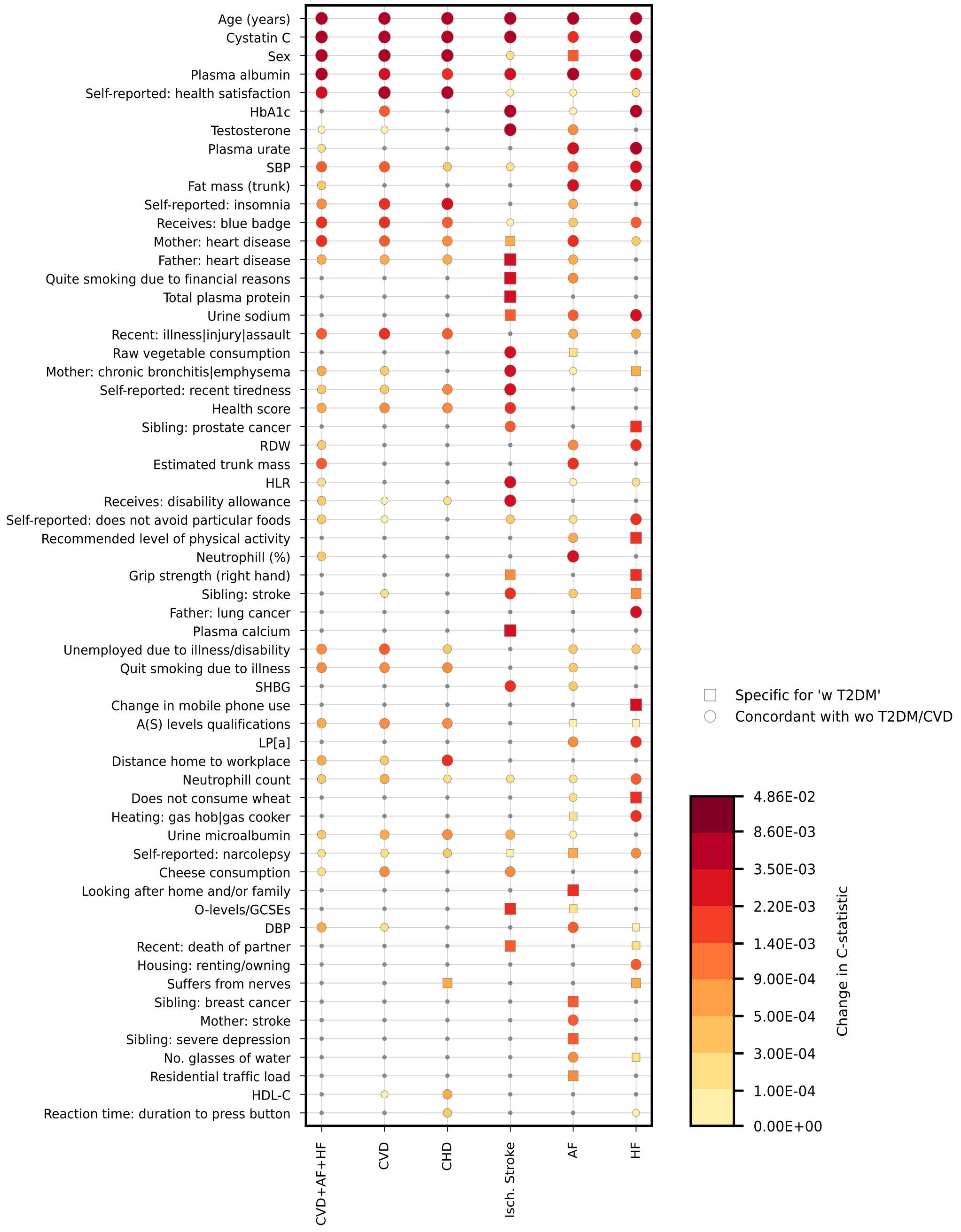

N. B. The y-axis presents the union of the top 20 features (limited to maximum 60 features) based on the c-statistic change for each of the six CVD outcomes. Plot markers (e.g. triangle, square) highlight the differences in identified features between groups. The permuted feature importance reflects the c-statistic change based on the test data; iteratively the values of each variable were randomly assigned to an individual after which the c-statistic was re-estimated with these permuted data and the difference in performance was used as an estimate of a variable contribution to the model's predictive potential. Abbreviations: glycated haemoglobin (HbA1c), systolic blood pressure (SBP), red blood cell distribution width (RDW), high light scatter reticulocyte count (HLR), sex hormone-binding globulin (SHBG), Qualifications (O levels / General Certificate of Secondary Education or equivalent) (O-levels/GCSEs), high-density lipoprotein cholesterol (HDL-C), diastolic blood pressure (DBP), lipoprotein A (LP[a]).

**Figure 6:** The contribution of the top 60 features to the prediction of six facets of CVD for ”w T2DM&CVD” group.

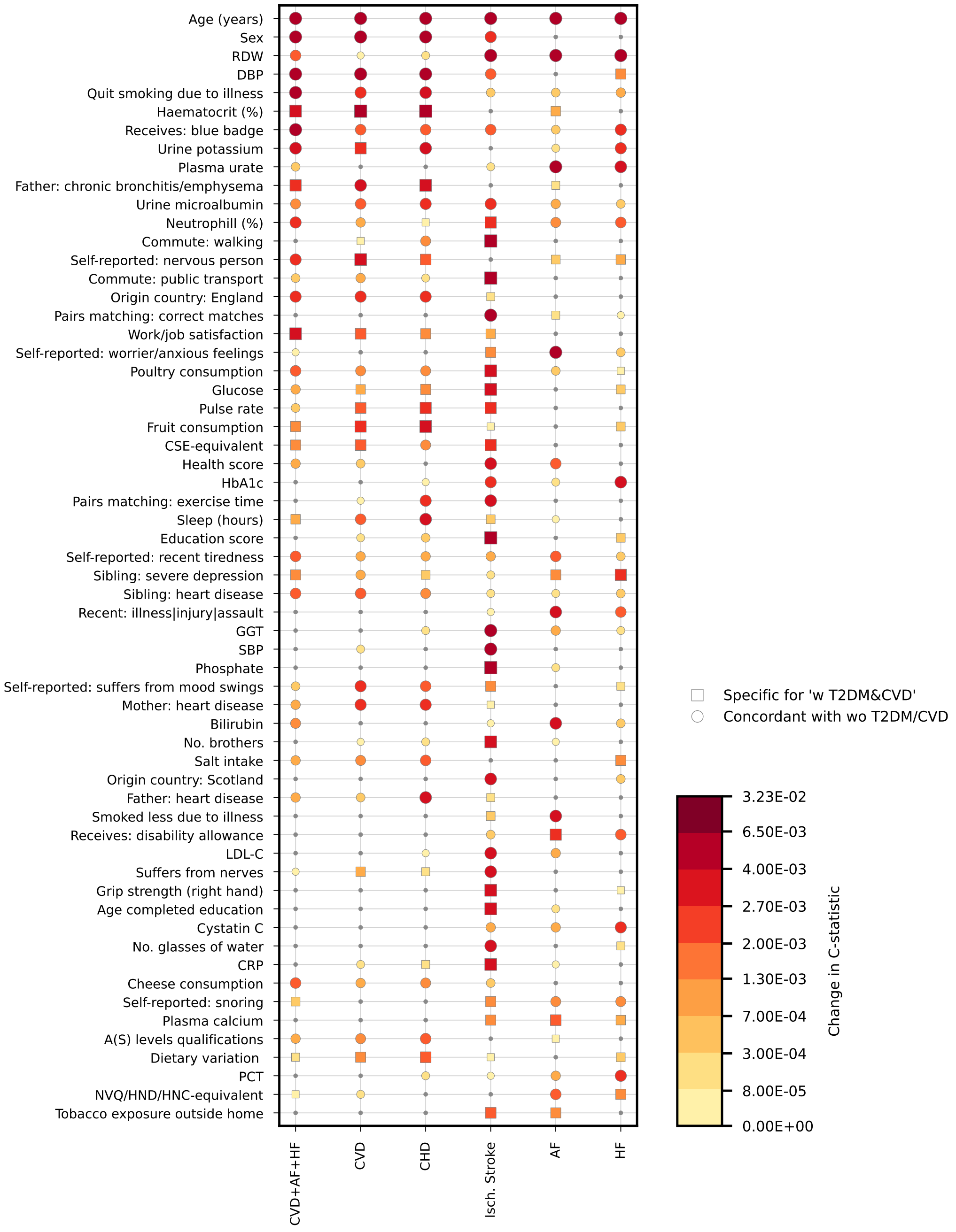

N. B. The y-axis presents the union of the top 20 features (limited to maximum 60 features) based on the c-statistic change for each of the six CVD outcomes. Plot markers (e.g. triangle, square) highlight the differences in identified features between groups. The permuted feature importance reflects the c-statistic change based on the test data; iteratively the values of each variable were randomly assigned to an individual after which the c-statistic was re-estimated with these permuted data and the difference in performance was used as an estimate of a variable contribution to the model's predictive potential. Abbreviations: red blood cell distribution width (RDW), alanine aminotransferase (ALT), low-density lipoprotein cholesterol (LDL-C), gamma glutamyltransferase (GGT), systolic blood pressure (SBP), qualifications (CSEs or equivalent) (CSE-equivalent), glycated haemoglobin (HbA1c), c-reactive protein (CRP), platelet crit (PCT), qualifications (NVQ or HND or HNC or equivalent) (NVQ/HND/HNC-equivalent).
